# International travel in times of the COVID-19 pandemic: Evidence from German school breaks

**DOI:** 10.1101/2021.02.19.21252062

**Authors:** Andreas Backhaus

**Author notes:** The views expressed in this paper are those of the author and do not necessarily reflect the views of the Federal Institute for Population Research. The author thanks Lukas Buchheim, Sebastian Klüsener, and Andreas Steinmayr for helpful comments and discussions.

## Abstract

The COVID-19 pandemic has triggered severe global restrictions on international travel with the intention of limiting the spread of SARS-CoV-2 across countries. This paper studies the effects of the partial relaxation of these travel restrictions in Europe during the summer months of 2020. It exploits the staggered start of the summer school breaks across German states as an exogenous shock to the travel opportunities of the population. While the school breaks also increased mobility within Germany, the event study regressions include disaggregated and time-varying controls for domestic mobility and local COVID-19-related restrictions. The resulting intention-to-treat effects of the relaxed travel restrictions show a significant and sizable increase of the COVID-19 incidence in German counties during the later weeks of the school breaks.

## 1 Introduction

Following the outbreak of the SARS-CoV-2 pandemic, Germany implemented a number of non-pharmaceutical interventions (NPIs) to slow down the spread of the virus and to prevent the German health care system from being overwhelmed. After a steep increase in infections in March and a peak in April, the number of new confirmed cases of COVID-19 infections in Germany dropped sharply in the subsequent weeks, similar to the patterns observed in other European countries.

The restrictions on cross-border movements and international travel constituted one of the most drastic and unprecedented NPIs, as they brought travel both outside and within Europe largely to a hold during the early months of the pandemic. However, Germany’s restrictions on intra-EU travel were considerably relaxed on June 15 (Deutsche Welle, 2020), as indicated by the vertical line marking calendar week 25 in Panel 1a of Figure 1, closely followed or preceded by other EU countries. Three weeks later, the weekly incidence of COVID-19 in Germany began to increase. Over the course of the following six weeks, it more than doubled; continuing its ascent after a brief plateau between calendar weeks 34 and 36.

**Figure 1:**
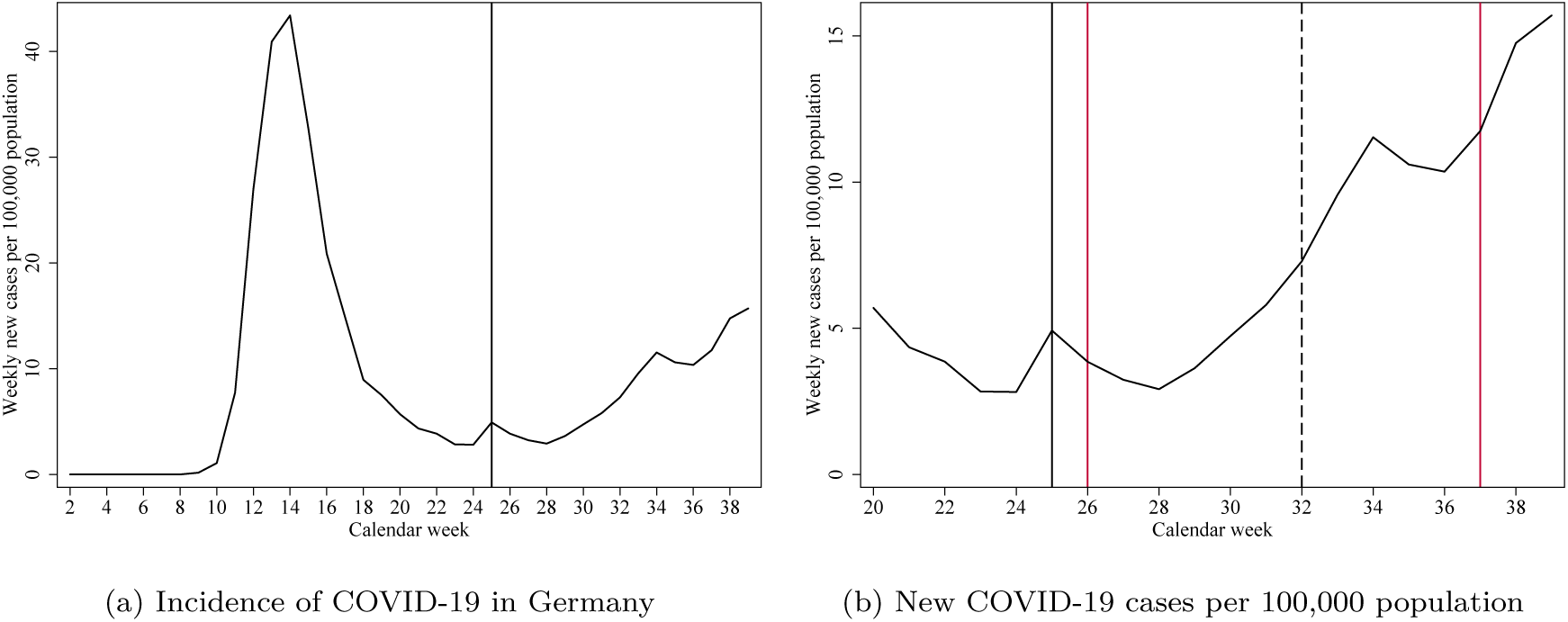
Incidence of COVID-19 and school breaks in Germany *Notes:* Source: RKI (2020). The left panel shows the weekly number of new confirmed cases of COVID-19 infections in Germany. Confirmed cases are expressed per 100,000 population. The vertical black line at week 25 indicates the relaxation of international travel restrictions. The right panel shows the weekly number of new confirmed cases of COVID-19 infections per 100,000 population in Germany. The vertical red lines at week 26 and week 37 indicate the earliest beginning and the latest conclusion of the school breaks in German states. The vertical dashed black line at week 32 indicates the beginning of a mandatory testing regime for returning travelers from risk areas as declared by the RKI.

This paper examines the relationship between the resurgence of international travel and the COVID-19 incidence in Germany. The empirical strategy exploits that the relaxation of the travel restrictions closely coincided with the beginning of the summer breaks in German schools. While the breaks generally last six weeks in all 16 German states, their timing is staggered across the summer months. The first states went on school breaks in calendar week 26, while the latest states concluded their school breaks not before calendar week 37. This period is indicated by the two vertical red lines in Panel 1b of Figure 1. The timing of the school breaks has further not been changed due to the pandemic. The exogenous and staggered timing of the school breaks therefore provides an ideal setting for an event study approach. The same setting has already been used by Isphording *et al*. (2020) and von Bismarck-Osten *et al*. (2020) to evaluate the effect of school closures and reopenings on the COVID-19 incidence in Germany. In the context of this study, the staggered school breaks represent an exogenous shock to the probability that individuals and families with school-aged children residing in a specific German state will travel during the summer months.

The hypothesis that international travel may have contributed to the rising COVID-19 incidence in Germany is motivated by the fact that residents of Germany traveled to other European countries that exhibited a considerably higher COVID-19 incidence during the summer months than Germany. Hence, at least at the national level, these travelers were exposed to environments that carried a relatively higher risk of infection, implying the potential for importing infections into Germany. The per-capita incidence of COVID-19 in several popular European summer travel destinations in comparison to the incidence in Germany is displayed in Figure 2, with the red vertical lines indicating the total duration of all school breaks in Germany. Most of the displayed countries had a much higher incidence than Germany, not even attempting to take into account that surveillance of COVID-19 may have been more constrained in some of these countries.

**Figure 2:**
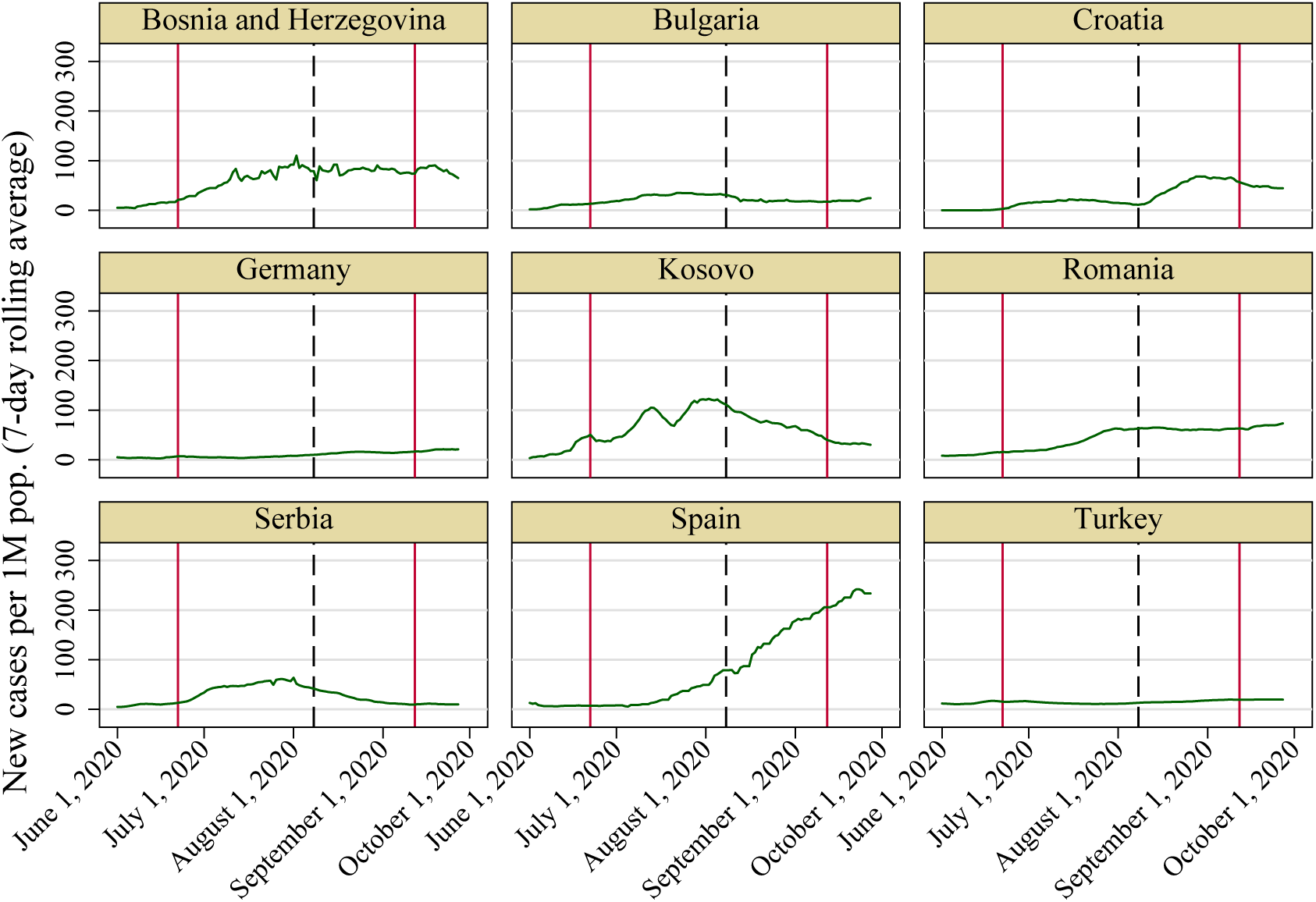
Cases in Germany and other European countries during the summer months of 2020 *Notes:* Source: Roser et al. (2020). The red vertical lines in each graph indicate the start of the earliest school break and the end of the latest school break respectively in Germany in 2020. The black dashed line in each graph indicates the implementation of the mandatory testing regime for returning travelers from risk areas.

The results of this study indicate a statistically significant increase in COVID-19 incidence in German counties in the second half of the school breaks. This pattern is robust to a variety of disaggregated and time-varying controls for mobility and COVID-19 restrictions within Germany. Magnitude and dynamics of the event study estimates are consistent with descriptive statistics on infections detected among travelers returning from abroad during the school breaks.

The paper contributes to the fast-growing evidence on the effectiveness of NPIs in containing the COVID-19 pandemic. A major advantage over several other studies is that the timing of the school breaks had been exogenously determined before the beginning of the pandemic. This feature diminishes the threat of reverse causality, which potentially affects the validity of studies examining the introduction of NPIs such as mandatory face mask mandates and stay-at-home orders, as NPIs are typically introduced when the epidemiological situation is demanding them. While overall, the relaxation of the travel restrictions was potentially only possible because of the relatively low incidence at that time, the particular shock to the probability of a state’s population to travel arrived exogenously via the staggered school breaks. A notable limitation of this paper, in turn, is that actual travel is unobserved both at the individual level and at higher levels of aggregation such as counties. The results should therefore be interpreted as intention-to-treat effects. Descriptive evidence on travel movements at the state- and national level during the summer months of 2020 is shown in support of the hypothesis that the loosened travel restrictions increased exposure to countries with a higher prevalence of COVID-19.

## 2 Literature review

Several studies have documented the role of international travel in the early spread of SARS-CoV-2 across countries (Zhang *et al*., 2020; Murphy *et al*., 2020; Böhmer *et al*., 2020; Rothe *et al*., 2020). A number of modeling studies have then attempted to assess the effectiveness of travel restrictions and border closures on the spread of the virus, e.g. Brady *et al*. (2020); Chinazzi *et al*. (2020); Wells *et al*. (2020); Costantino *et al*. (2020); Wu *et al*. (2020); Linka *et al*. (2020); Ruktanonchai *et al*. (2020); Russell *et al*. (2020). Another strand of the literature has estimated the effectiveness of border closures and travel restrictions without relying on epidemiological models (Koh *et al*., 2020; Kraemer *et al*., 2020; Keita, 2020; Eckardt *et al*., 2020). Typically, these studies examine a bundle of NPIs enacted during the early stage of the pandemic. However, the close succession of NPIs and the uncertainty surrounding the accurate surveillance of SARS-CoV-2 in this period present two challenges to their approach. Further, these studies typically cannot address the potential reverse causality between the enactment of NPIs and the epidemiological situation.

A meta-review on travel-related control measures published in September 2020, noting the imbalance between modeling studies and observational studies, assessed a lack of ‘real-life’ evidence on the effectiveness of these measures (Burns *et al*., 2020). The certainty of the evidence for most travel-related control measures was rated as low, due to inappropriate assumptions in the modeling studies on the one hand and potential bias in the observational studies on the other hand. Hence, it is reasonable to complement the existing literature with evidence based on exogenous variation in travel opportunities.

## 3 Context

### 3.1 Restrictions on international travel during the pandemic

Restrictions on international travel were imposed globally in the early months of the pandemic in order to prevent and limit the transmissions of the virus across national borders. From a European perspective, the restrictions affected travel both between EU countries on the one hand and between the EU, other European countries, and the rest of the world on the other hand. In mid-March, the EU closed its external borders for travelers from non-EU countries (Deutsche Welle, 2020b). Simultaneously, the EU member states put in place restrictions on non-essential travel of EU citizens across their national borders, with limited exceptions. The EU member states differed in the detailed design of the travel restrictions applying to their respective territories. Germany issued a global travel warning in mid-March, advising its citizens against any travel abroad, while imposing border controls at its borders to neighboring countries (Deutsche Welle, 2020a).

*De facto*, the coordinated actions taken by the EU member states severely halted both mobility within the EU and mobility between the EU and the rest of the world. The magnitude of the downturn in international mobility from and to Germany can be exemplified by statistics on commercial air travel. Figure 3 indicates a sharp decrease in flight movements in the German airspace beginning in early March (calendar weeks 10/11), with flight movements hitting a tentative yearly low in early April 2020 (calendar week 15/16). Around this time, air traffic including military and non-passenger flights was more than 80% lower than in the same weeks of 2019.

**Figure 3:**
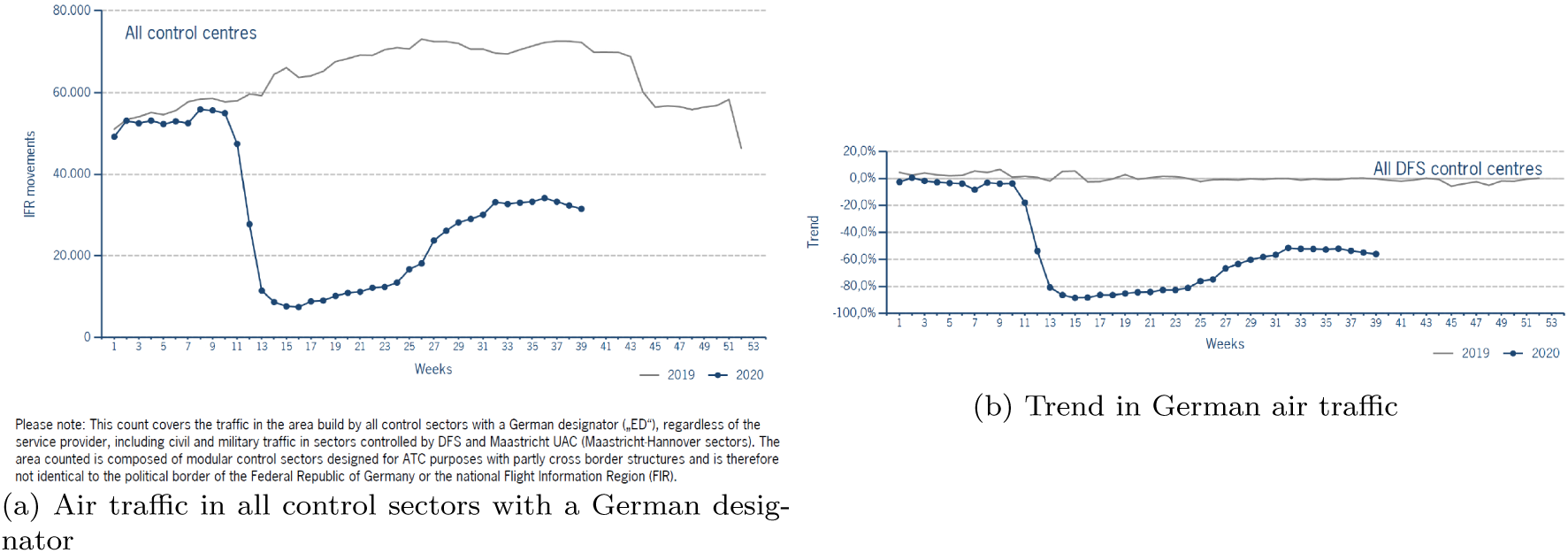
Air traffic in Germany during the COVID-19 pandemic *Notes:* DFS (2020). The left panel shows the absolute weekly number of movements in the area build by all control sectors with a German designator, regardless of the service provider, including civil and military traffic in sectors controlled by DFS and Maastricht UAC (Maastricht-Hannover sectors). The right panel shows the weekly deviation of traffic in 2020 from the 2019 level in the area build by control sectors of DFS control centres, including civil and military traffic. Traffic in Maastricht UAC (Maastricht-Hannover sectors) is not counted.

The travel restrictions remained in place till the summer months regarding travel within Europe. By mid-June then, the EU made coordinated efforts to revive travel between its member states and nearby countries (Deutsche Welle, 2020). Complementary, the German government revoked its travel warnings for most of the other EU countries. This policy change is mirrored by the accelerated recovery of the air traffic from and to Germany from calendar week 25 onward, as shown in Figure 3. The German federal government maintained surveillance of the epidemiological situation in the rest of Europe via the RKI^1^ by implementing a two-step procedure for issuing new travel warnings: First, countries or sub-national regions would be identified where the 7-day-incidence of COVID-19 exceeded the level of 50 new cases per 100,000 population. Then, additional qualitative criteria are used to determine whether or not countries or regions that might nominally fall below this threshold could nonetheless still pose an increased risk of infection; with the same rationale applying to countries or regions that might nominally fall above this threshold but do not nonetheless pose an increased risk (Robert Koch-Institut, 2021b). After consultation of the responsible ministries and agencies, such places would then be designated as ‘risk areas’. Consequently, travel warnings were issued for a number of European countries and for most countries in the rest of the world simultaneously with the general relaxation of the travel restrictions.

Transportation statistics suggest that the revived air travel from and to Germany remained largely focused on Europe during the summer months: More than 90% of passengers who departed from German airports during the four months from June to September 2020 had other European countries as their flights’ countries of destination; similarly, 90% of passengers arriving at German airports during this period arrived from other European countries (Statistisches Bundesamt (Destatis), 2020b).

However, while travel remained largely restricted to Europe, the travel warnings maintained or reissued by the German federal government do not appear to have regulated travel between Germany and other European countries during the summer months of 2020. For example, the travel warnings for the Southeast European states of Kosovo and Serbia were never rescinded during this period; nonetheless, more than 95,000 passengers departed from German airports towards these two countries between June and September 2020 (Robert Koch-Institut, 2020b; Statistisches Bundesamt (Destatis), 2020b). Furthermore, travel warnings were only rescinded for some regions of Turkey in early August; however, the number of passengers departing towards Turkey had already surged in June and remained high throughout the summer. In addition, some countries have only been (re-)designated as risk areas nationwide after cases had already been on the rise there for some time: Spain, for example, with the exception of the Canary Islands, was re-designated as a risk area on August 14, while the country had already crossed the 7-day-incidence threshold of 50 new cases per 100,000 population on August 3 (Robert Koch-Institut, 2020a; Roser *et al*., 2020). Exemplary, Figure 4 displays the number of passengers departing from Germany to states in Southeast Europe, Turkey, and Spain over the first nine months of 2020.

**Figure 4:**
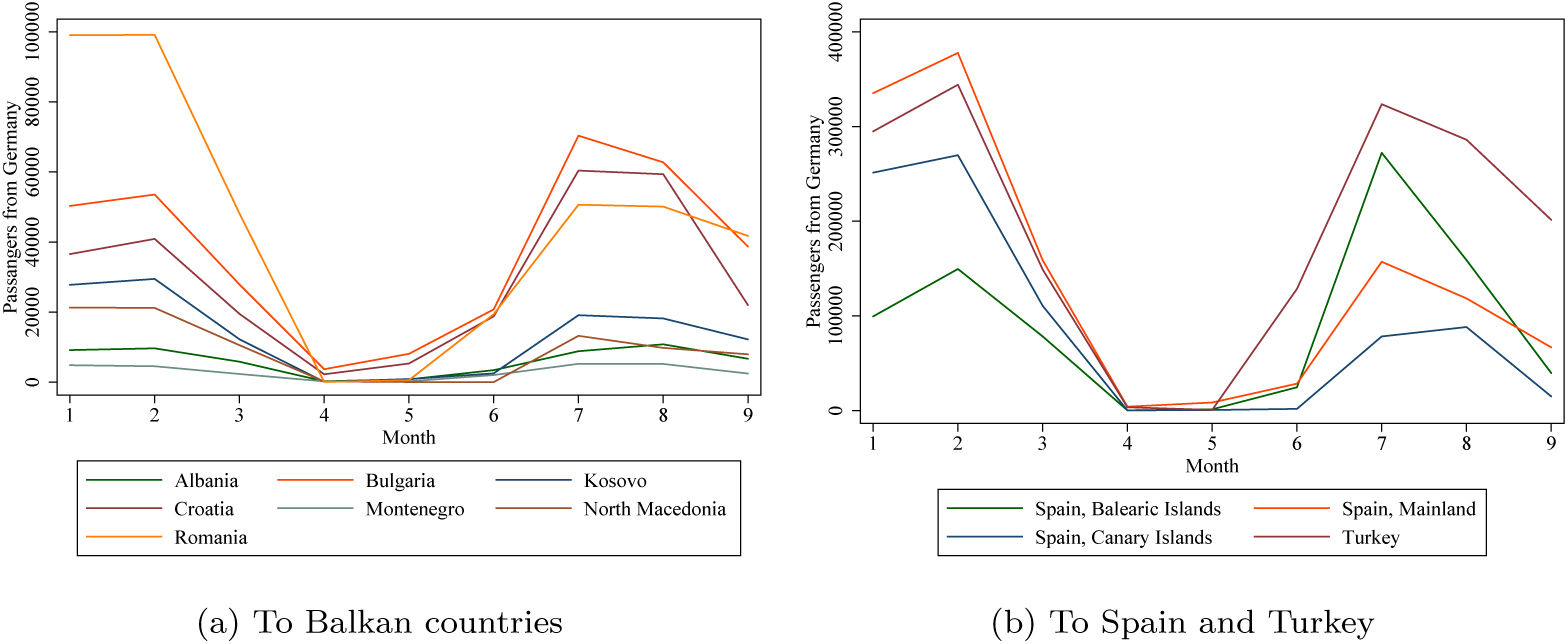
Passengers departing from German airports during the summer months of 2020 *Notes:* Destatis (2020). Panel 4a displays the number of passengers departing from airport in Germany to a number of Southeast European countries during the first nine months of 2020. Panel 4b displays the number of passengers departing from airport in Germany to Turkey and several regions of Spain during the first nine months of 2020.

### 3.2 Epidemiological situation of travel returnees

Over the course of the summer months, evidence began to accumulate at local public health office and the RKI that a growing number of infections confirmed by tests in Germany had been contracted abroad. This tendency is reflected in the share of confirmed cases in Germany with the probable place of infection abroad displayed in Figure 5: Following the relaxation of the travel restrictions in calendar week 25, this share first rose slowly to approx. 10% but then surged to approx. 50% between week 30 and week 34. Following its peak in mid-August, the share of confirmed cases with probable infection abroad began to decline again, first slowly, then rapidly from week 36 onward, reverting back to a level of approx. 10% in week 39 (September 21-29). Simultaneously, the confirmed cases with probable infection abroad also declined strongly in absolute terms.

**Figure 5:**
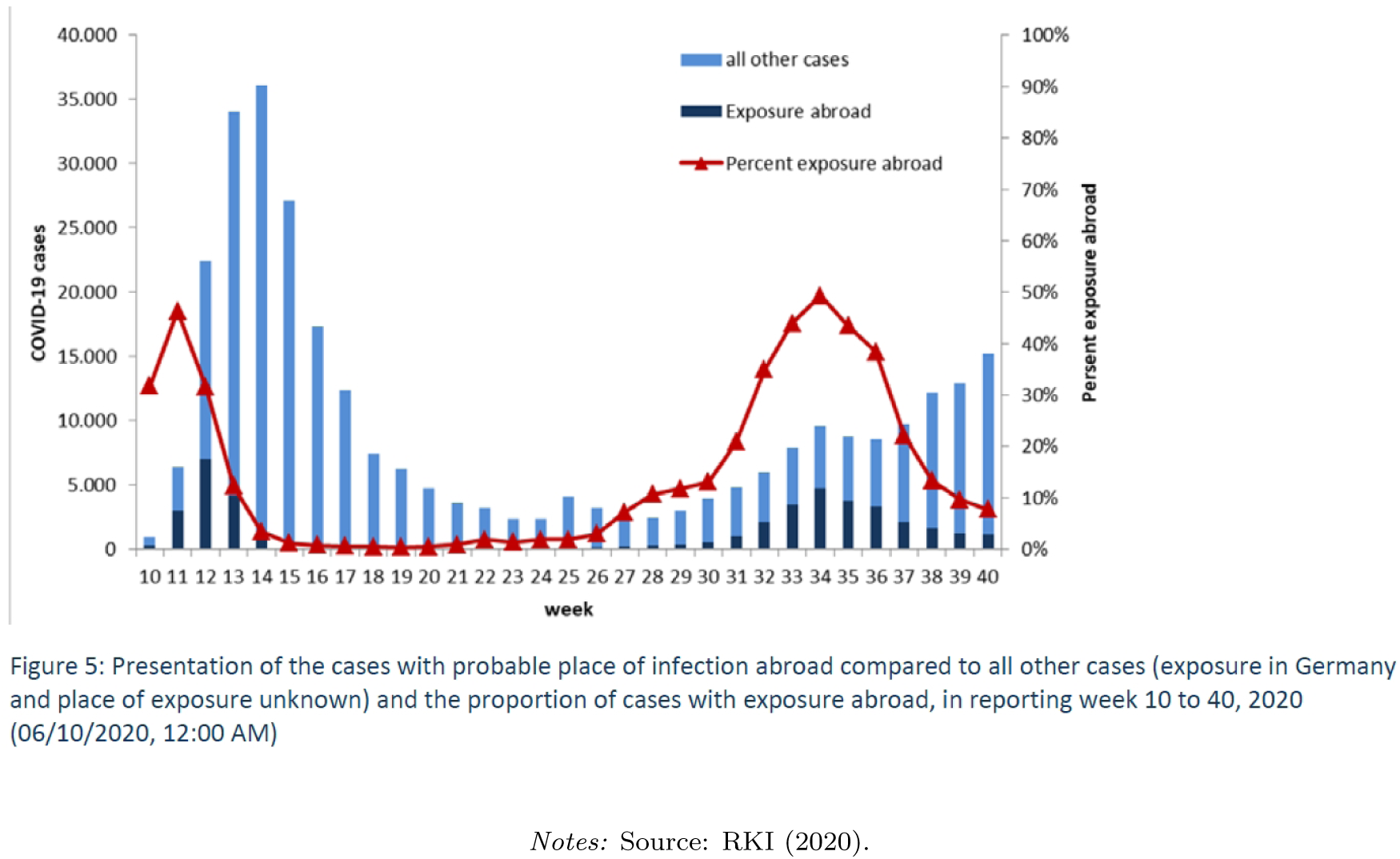
Cases in Germany with exposure abroad

This surge in cases with probable infection abroad was partly accompanied by the introduction of a free testing regime for all returning travelers from non-risk areas on August 1 and a mandatory free testing regime for travelers returning from the designated risk areas on August 8. While travelers returning from risk areas had already been required to quarantine for 14 days in their residences in accordance with regulations passed by the German states in mid-July, enforcement of the quarantine was in the hands of the local public health offices, relying at least partly on voluntary compliance. The free and voluntary tests for returnees in general and the free and mandatory tests for returnees from risk areas were hence suitable for improving the surveillance of infections among returnees. While the free voluntary testing regime was terminated on September 15, or three days after the last state had completed its school breaks, the mandatory testing regime remained in place. (Deutsche Welle, 2020e) The potential impact of the two testing regimes on the estimation results of this study is discussed in Section 7.

In total, the RKI registered more than 24,000 confirmed cases with likely place of infection abroad among returning travelers between the calendar weeks 26 to 39 (06/22/2020-09/27/2020) (Robert Koch-Institut, 2020a). This figure provides a useful orientation regarding the number of infections introduced into Germany from returning travelers during the summer months. Table 1 highlights that only five travel destination countries account for almost 14,000 of the 24,000 confirmed cases among travel returnees, with three of the five countries being located in Southeast Europe.

**Table 1:** Number of infected travel returnees by country of likely infection and calendar week

| Week | Kosovo | Croatia | Turkey | Romania | Spain |
| --- | --- | --- | --- | --- | --- |
| 27-30 | 303 | 29 | 70 | 36 | 17 |
| 31 | 341 | 45 | 123 | 40 | 27 |
| 32 | 564 | 235 | 393 | 56 | 76 |
| 33 | 847 | 588 | 670 | 111 | 120 |
| 34 | 958 | 1153 | 496 | 174 | 296 |
| 35 | 767 | 895 | 363 | 208 | 211 |
| 36 | 426 | 638 | 403 | 208 | 140 |
| 37 | 168 | 208 | 346 | 165 | 103 |
| 38 | 91 | 90 | 261 | 103 | 60 |
| 39 | 44 | 43 | 156 | 84 | 46 |
| Total | 4509 | 3924 | 3281 | 1185 | 1096 |

*Notes:* Source: RKI Situation Reports.

However, not all cases of infected travel returnees detected in Germany may have been correctly classified as having their origin of infection abroad. Further, infected travel returnees may have caused secondary infections and subsequently detected cases among their contacts back in Germany. In addition, travel to destinations abroad had already been picking up pace since June, as indicated in Section 3.1, and by early August, all states had already begun their school breaks, with some concluding them shortly before or around the time when the free and mandatory testing regimes were implemented. Taken together, there is reason to suspect that the public surveillance of the travel returnees may not have fully captured the contribution of international travel to the epidemiological situation in Germany during the summer months of 2020.

### 3.3 Summer school breaks in Germany

The empirical strategy of this paper exploits the staggered timing of school summer vacations across Germany’s 16 federal states as an exogenous shock to the opportunities of the population to embark on international travels. While the summer break lasts six weeks in every state, the start and end dates of the break generally differ in each year depending on the state of residency. The start and end dates for the 2020 school summer vacations were set before the beginning of the pandemic; they have not been altered since then. The earliest state to begin the summer break was Mecklenburg-Vorpommern on June 22 in calendar week 26, shortly after the suspension of the travel warnings, while the latest state to conclude the summer break was Baden-Württemberg on September 12 in calendar week 37. Hence, schools have been on the six-week summer break in at least one German state over a total period of twelve consecutive weeks. All start and end dates in the respective states are displayed in Table 2.

**Table 2:**
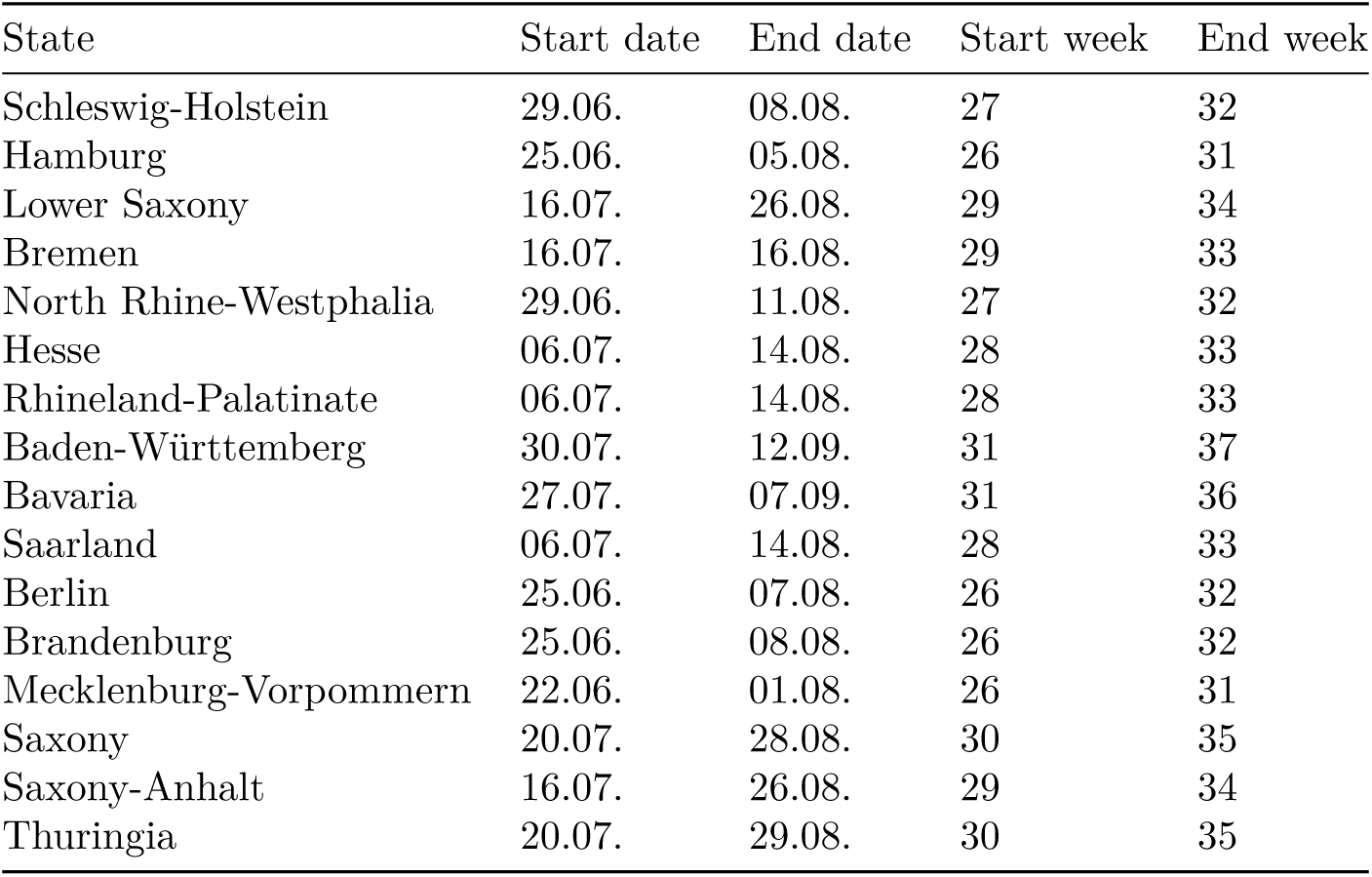
Start and end dates of summer school breaks in German states

It is reasonable to assume that the duration of the school breaks served as a stern constraint on the ability of families with school-aged children to travel. While schools had been closed during the initial months of the pandemic in Germany, the states had returned to at least partial or rotating in-class instruction before the start of the school breaks. Further, return to in-class instruction had been announced and was henceforth expected after the summer break. Therefor, travel with school-aged children could not be delayed until after the end of the school breaks. Finally, even if significant travel with school-aged children already occurred before the official start of the school breaks, this would bias the estimate of the school break effects towards zero, as travel and potential imported cases would occur earlier than the official dates of the school vacations would suggest.

Regarding the relative size of the population whose travel opportunities were affected by the school breaks, it can be said that families with underaged children represent at least 30% of the population in every German state, as displayed in Table 3. The mobility potential of the population in question is therefore sufficiently large to affect the general population incidence.

**Table 3:** Share of family members among the population by German states

| State | Share of family members among population |
| --- | --- |
| Schleswig-Holstein | 0.3464 |
| Hamburg | 0.3627 |
| Niedersachsen | 0.3569 |
| Bremen | 0.3347 |
| Nordrhein-Westfalen | 0.3627 |
| Hessen | 0.3680 |
| Rheinland-Pfalz | 0.3576 |
| Baden-Württemberg | 0.3715 |
| Bayern | 0.3626 |
| Saarland | 0.3253 |
| Berlin | 0.3499 |
| Brandenburg | 0.3418 |
| Mecklenburg-Vorpommern | 0.3265 |
| Sachsen | 0.3374 |
| Sachsen-Anhalt | 0.3144 |
| Thüringen | 0.3262 |

*Notes:* Source: Federal Statistical Office. The table reports the share of family members among the population in each of Germany's states in 2019. Only families with children younger than 18 years are counted.

Descriptive evidence on the association between school breaks and travel behavior is presented in Figure 6. The graph displays the number of passengers departing from five major German airports to destinations abroad over the course of the summer months. The airports Berlin-Tegel, Düsseldorf, and Hamburg are located in states that went on school breaks before July, while the airports München and Stuttgart are located in states that went on school breaks near the end of July. Due to its proximity to neighboring states, Frankfurt airport is not included in the graph. Notably, the number of passengers departing from Berlin-Tegel, Düsseldorf, and Hamburg already peaked in the month of July, while it peaked one month later in München and Stuttgart. This pattern is consistent with the hypothesis that the earlier school breaks in Berlin, North Rhine-Westphalia, and Hamburg caused an earlier recovery of international departures from these states.

**Figure 6:**
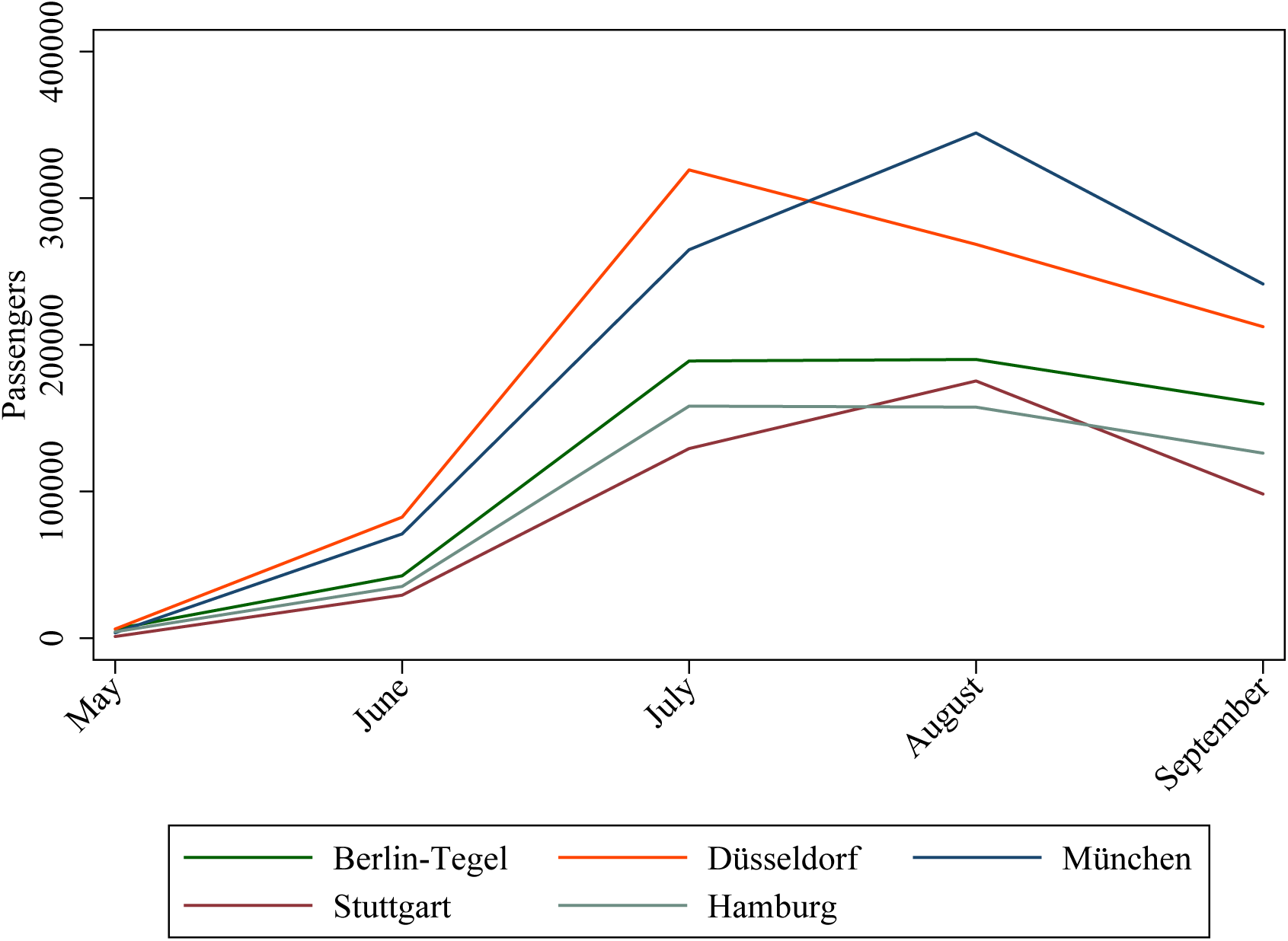
Passengers departing from German airports during the summer months of 2020 *Notes:* Source: Destatis (2020). Each line represents the number of passengers departing from the indicated German airport in the indicated month towards destinations abroad.

Furthermore, Figure 7 displays descriptive evidence on the evolution of the weekly COVID-19 incidence per 100,000 population for each German state. In each and every state, the weekly incidence of COVID-19 was higher by the end of the school breaks than by their beginning. The three most populous states, Baden-Württemberg, Bavaria and North Rhine-Westphalia, have all seen strong increases in their COVID-19 incidence.

**Figure 7:**
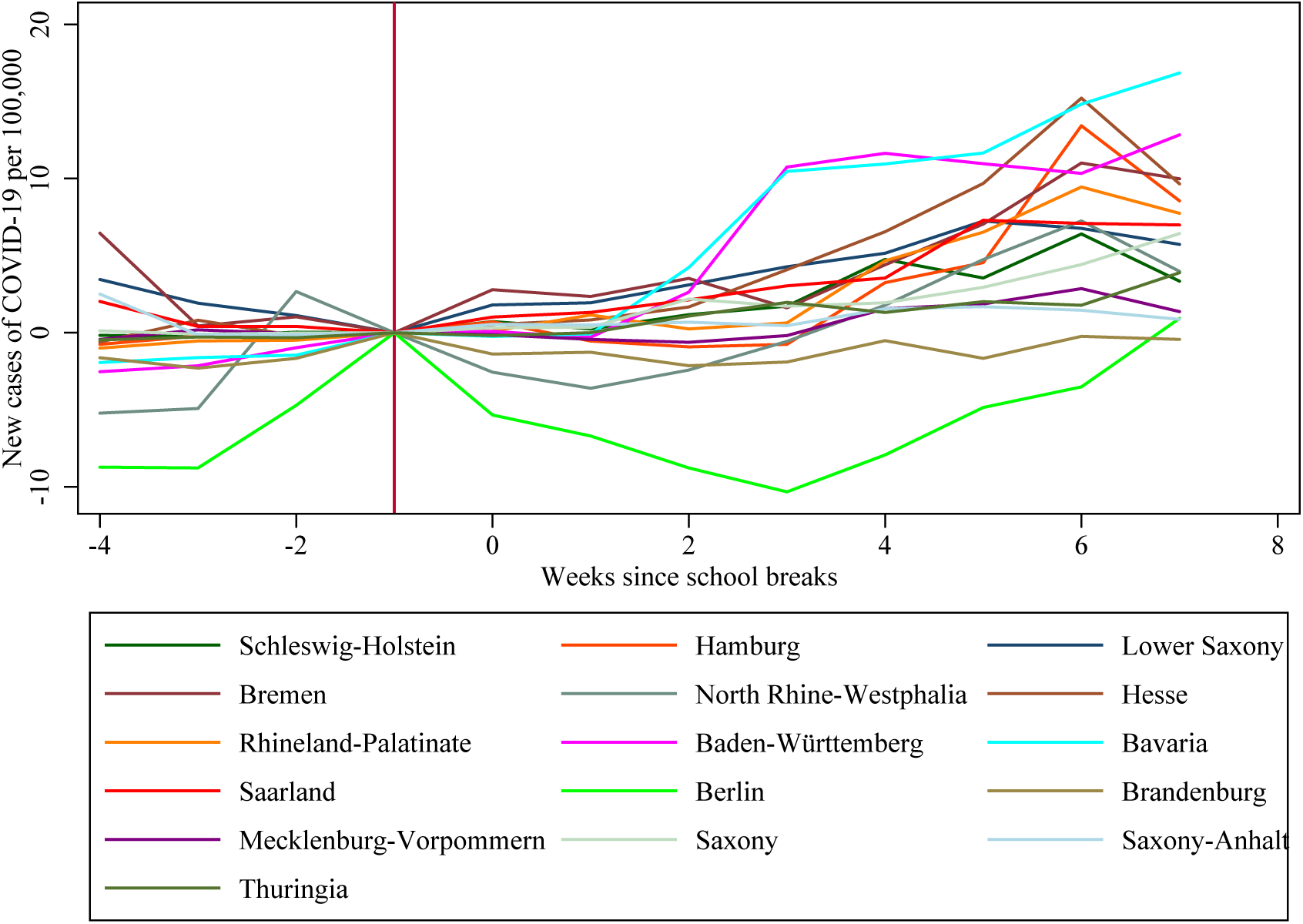
New cases of COVID-19 by state before and during the school breaks *Notes:* The graph displays the evolution of new confirmed cases of COVID-19 per 100,000 population in each of Germany’s 16 states. New cases are displayed up to four weeks before and seven weeks since the start of the school breaks in each state. The last period before the beginning of the school breaks is used as the reference period.

## 4 Empirical strategy

The empirical investigation into whether international travel increased the COVID-19 incidence in Germany is complicated by the fact that the travel restrictions were relaxed simultaneously all across Germany, thereby not providing a control group of German locations where the restrictions were still in place. However, while the international travel restrictions were eased simultaneously all over Germany, the staggered starting dates of the school breaks across states provide exogenous variation in the opportunities to travel, primarily for families with school-aged children. At the beginning of the observation period, all units are untreated, meaning no state is on school break yet. Over time, more and more states start their school breaks and are hence treated. After a certain date, all states have begun their school breaks, which corresponds to the treatment being switched on for all units in the sample. The setting of the German summer school breaks therefore presents the opportunity for an event study approach.

The event study design is particularly convenient for the case of international travel following the start of the school breaks, as it is not *a priori* obvious when the treatment could show an effect: Following the beginning of the school breaks, people have to travel, they have to get infected, and they have to return while still infected or having overcome their infection only recently in order to test positive upon or after arrival back in Germany. In the event study design, the treatment effects are allowed to vary over time; hence the corresponding regression model will yield an individual estimate for each period both before and after the beginning of the school breaks. By contrast, estimates would be biased away from the true treatment effects in a DiD design with two-way fixed effects, staggered treatment, and varying treatment effects over time (Goodman-Bacon and Marcus, 2020).

The exogenous timing of the school breaks further alleviates concerns in many other studies evaluating the effects of NPIs, namely that a specific restriction or a package thereof would be introduced (or lifted) more likely in places that necessitate (or allow) it. While overall, the lifting of travel restrictions was certainly encouraged by the low COVID-19 incidence in early June, the beginning and the end of the school breaks in the various states were unaffected by the epidemiological situation.

The obvious threat to this identification strategy is the possibility that the school breaks may have increased COVID-19 incidence in Germany not only by inducing more international travel and hence the introduction of infections from abroad, but also by increasing mobility and social contacts within Germany, thus increasing COVID-19 incidence without any significant contribution from international travel. In addition, COVID-19 restrictions unrelated to international travel have become more scattered across the German states in terms of their strictness as compared to the first nationwide contact restrictions in spring.

Figure 8 displays graphs of various state-level Google mobility indicators before and since the start of the school breaks. Relative to a baseline period between January and February 2020, the graphs indicate a strong increase in mobility only for the states of Mecklenburg-Vorpommern, Schleswig-Holstein, and Brandenburg. However, as indicated in Figure 7, these states experienced a very low COVID-19 incidence at the same time. Further, while the frequency of visits to parks will naturally be at a higher level during summer than during winter, the risk of infection in open spaces is considered to be relatively low. Complementary, Figure 9 displays the state-level mobility before and since the start of the school breaks as measured by mobile phone data relative to the previous year. In accordance with the Google mobility data, the three aforementioned states experience the strongest increase in mobility.

**Figure 8:**
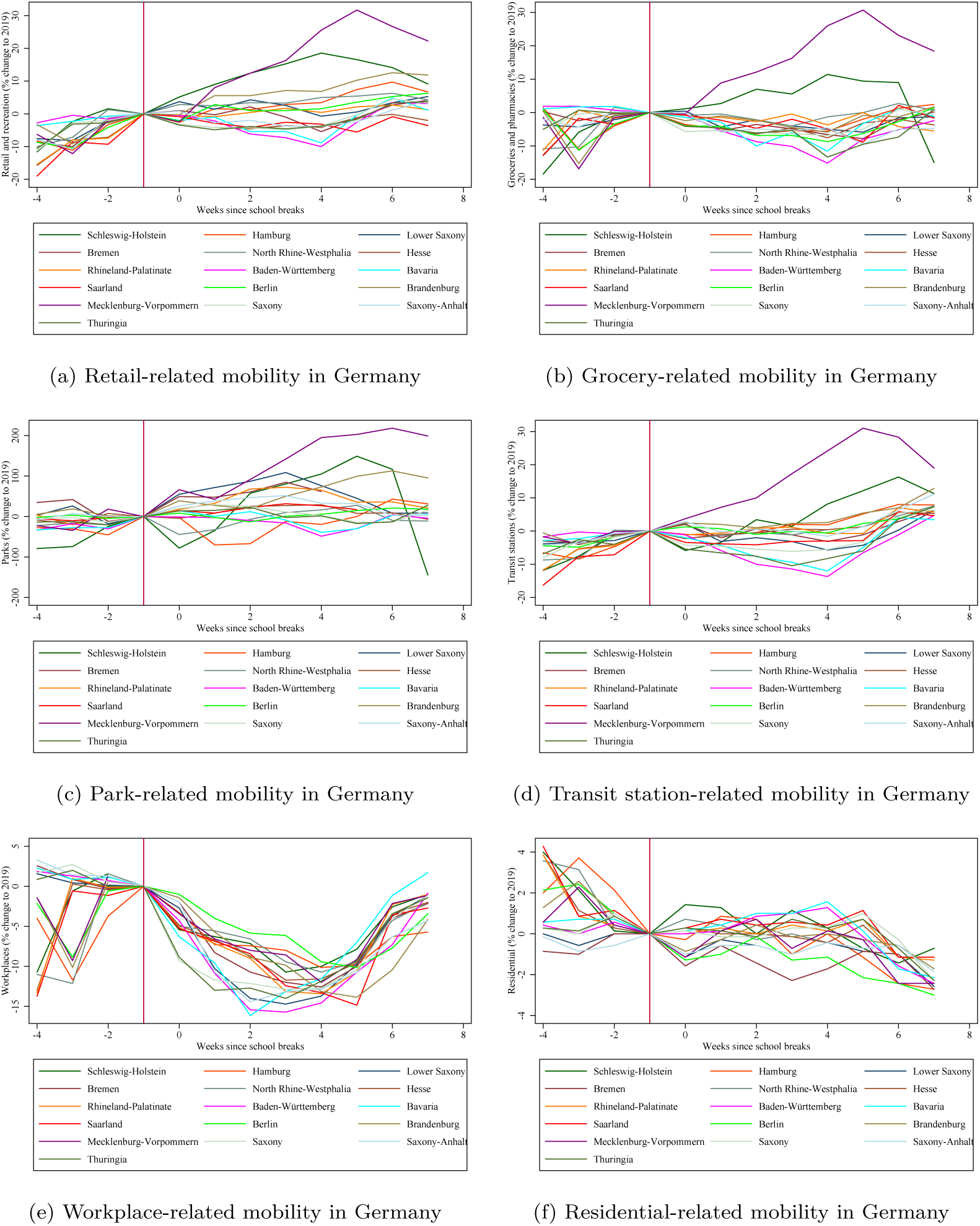
Google mobility trends in Germany before and during the school breaks *Notes:* Source: Google LLC (2020). Each panel displays the evolution of a different Google mobility indicator during the weeks before and after the beginning of the school breaks in 2020. Each line indicates a different German state. The last period before the beginning of the school breaks is used as the reference period.

**Figure 9:**
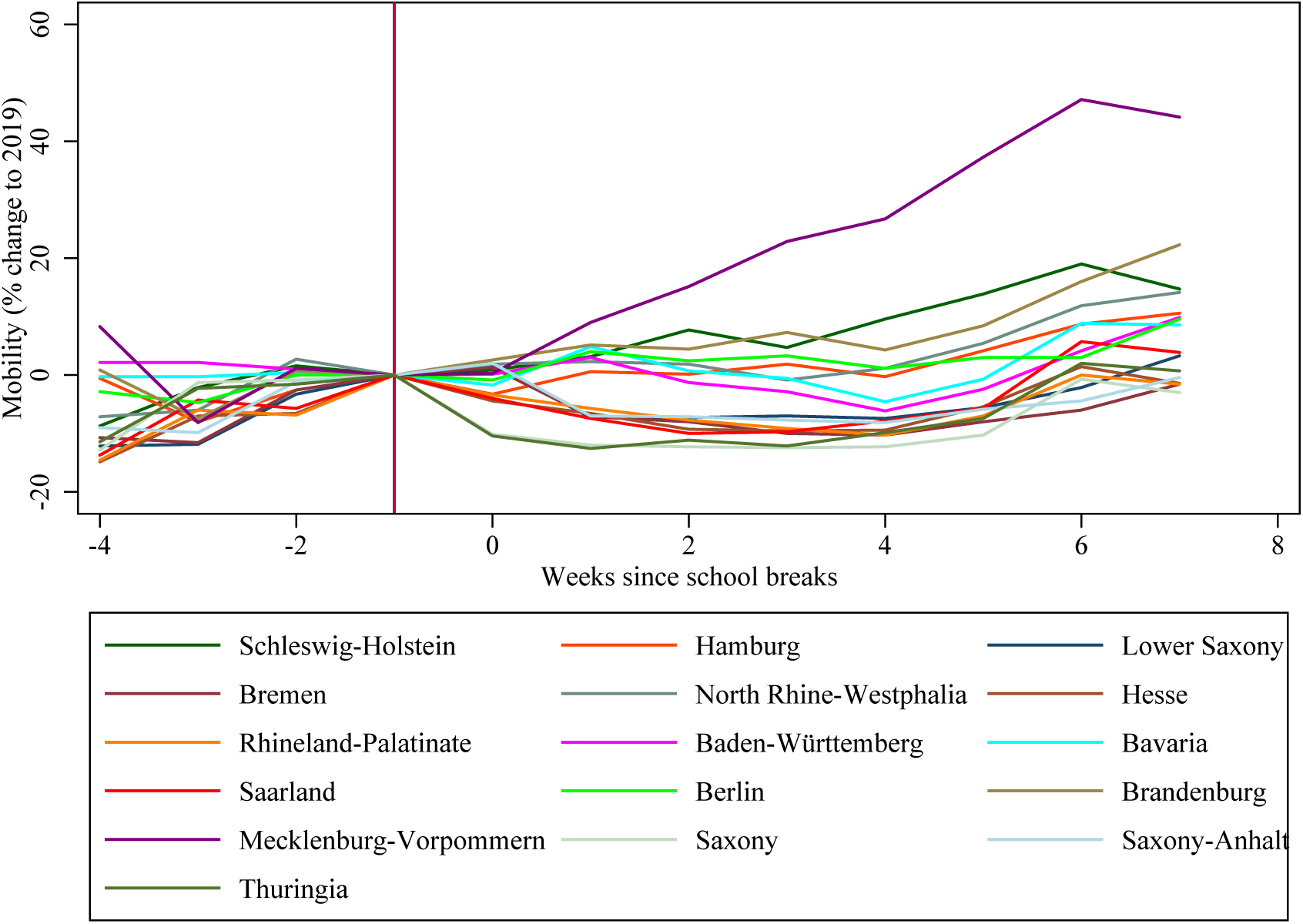
Mobility trends in Germany before and during the school breaks *Notes:* Source: Destatis (2020). The graph displays the evolution of mobility in each of Germany’s 16 states before and after the beginning of the school breaks in 2020. The last period before the beginning of the school breaks is used as the reference period.

Still, it is indispensable to control for changes in mobility and restrictions over time and at a more disaggregated level. Fortunately, data are available to construct such time-varying controls at the county level for this study’s period of interest.

Given that actual travel is unobserved, the effect of the school breaks on new cases of COVID-19 is estimated by directly regressing the new cases in Germany on the set of event indicators. The results should therefore interpreted as intention-to-treat (ITT) effects, as not every resident in Germany experienced a shock to her/his opportunities to travel from the combination of the relaxed restrictions and the onset of the school breaks - among those who have, not everyone has actually traveled.

Following the notation proposed by Clarke and Schythe (2020b), the regression model for the event study is formulated as follows:

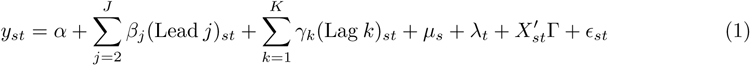

*y_st_* is the number of new confirmed cases of COVID-19 per 100,000 population in county *s* in week *t*. *α* is a constant. *µ_s_* and *λ_t_* are county and week fixed effects respectively, while *E_st_* is an unobserved error term. *X_st_* contains county-level and time-varying controls, which will capture patterns of mobility and changes to COVID-19-related restrictions during the observation period. The *J* leads and *K* lags are binary variables that indicate that the given state was a given number of periods away from the beginning of the school break in the respective time period. One period serves as baseline and is hence omitted; this is the last period before the onset of treatment, which corresponds to the first lead.

The main specification uses up to four leads and up to six lags. The rationale for using up to four lags is that this time window should be sufficient for detecting potential violations of the parallel trends assumption while not running the risk of picking up dynamics from Germany’s first wave. Statistically significant estimates of the *β_j_* coefficients would hence indicate a potential violation of the parallel trends assumption. In turn, statistically significant estimates of the *γ_k_* coefficients would indicate effects of the treatment, in this case the school breaks, on the COVID-19 incidence. While the school breaks last only six weeks, the sixth lead is intended to capture potential infections that have taken place in the last week of the school breaks but which have only been confirmed by test results in the following week. All regressions in the following are weighted by county population.

The regression results in this paper and their graphical representations are obtained by applying the user-written Stata routine eventdd (Clarke and Schythe, 2020a). Among its numerous functionalities, which are presented in more detail by Clarke and Schythe (2020b), this routine allows binning lead and lag periods beyond the specified maximum lead and lag periods into the final lead and lag terms, as suggested by Schmidheiny and Siegloch (2019) in settings where all units are eventually treated. This binning procedure is applied in all regressions reported in the following. The routine further provides the option to calculate confidence intervals by applying the wild cluster bootstrap method via the user-written Stata routine boottest (Roodman, 2015; Roodman *et al*., 2019). The wild cluster bootstrap is particularly relevant in the setting of this study: the timing of the summer breaks varies at the level of Germany’s states, which suggests clustering the standard errors of the coefficients at the state level. However, there are only 16 states in Germany, which may downward-bias the cluster robust variance estimate (Cameron and Miller, 2015). The wild cluster bootstrap has been shown to provide reliable inference even if the number of clusters is small (Cameron *et al*., 2008).

## 5 Data

Data on COVID-19 incidence in Germany are provided by Robert Koch-Institut (2020c). The data are available in daily format and disaggregated to the level of German counties (dt. *Kreise*). The daily reported cases are subject to considerable reporting variability across the days of the week due to some counties’ public health offices not reporting new cases on weekends. Therefore, the weekly instead of the daily incidence of COVID-19 per 100,000 population is computed and used as the dependent variable in the following. If school breaks do not start at the first day of the week in a given state, the state is defined as being on school break if the majority of weekdays in a given week fall into the break period.

The incidence dataset contains the date (dt. *Meldedatum*) when a new confirmed case of COVID-19 was reported to the RKI. The report to the RKI is usually the consequence of a positive result of a PCR test for SARS-CoV-2. Given that Germany has preferably been testing symptomatic individuals, a time lag of several days may exist between the infection date and the reporting date to the RKI, with the length of the lag depending on the length of the period between infection and the development of symptoms on the one hand, and on the length of the period between taking a test and receiving the result on the other hand. Given the rather large uncertainty surrounding this lag, the baseline regressions reported in the following do not make any adjustments in this regard but take the reported incidence as given.

County-level population data to compute the COVID-19 incidence per 100,000 population are provided by Statistisches Bundesamt (Destatis) (2020a). While not part of the main dataset, data collected by Roser *et al*. (2020) has been used in various graphical representations in this study. County-level mobility controls and controls for the level of COVID-19 related restriction are computed from data provided by Statistisches Bundesamt (Destatis) (2020c) and Infas 360 (2020). State-level mobility controls are computed from Google mobility data (Google LLC, 2020).

## 6 Results

### 6.1 Main results

Figure 10 displays the main results of this study. Each graph shows a plot of the estimated coefficients of the leads and lags. The horizontal axis indicates the weeks before resp. since the beginning of the summer school breaks. The first lead representing the baseline period is omitted. The 0 period hence indicates the week in which the event of the school breaks occurred for the first time. The vertical axis indicates the weekly incidence of new COVID-19 cases per 100,000 population in German counties.

**Figure 10:**
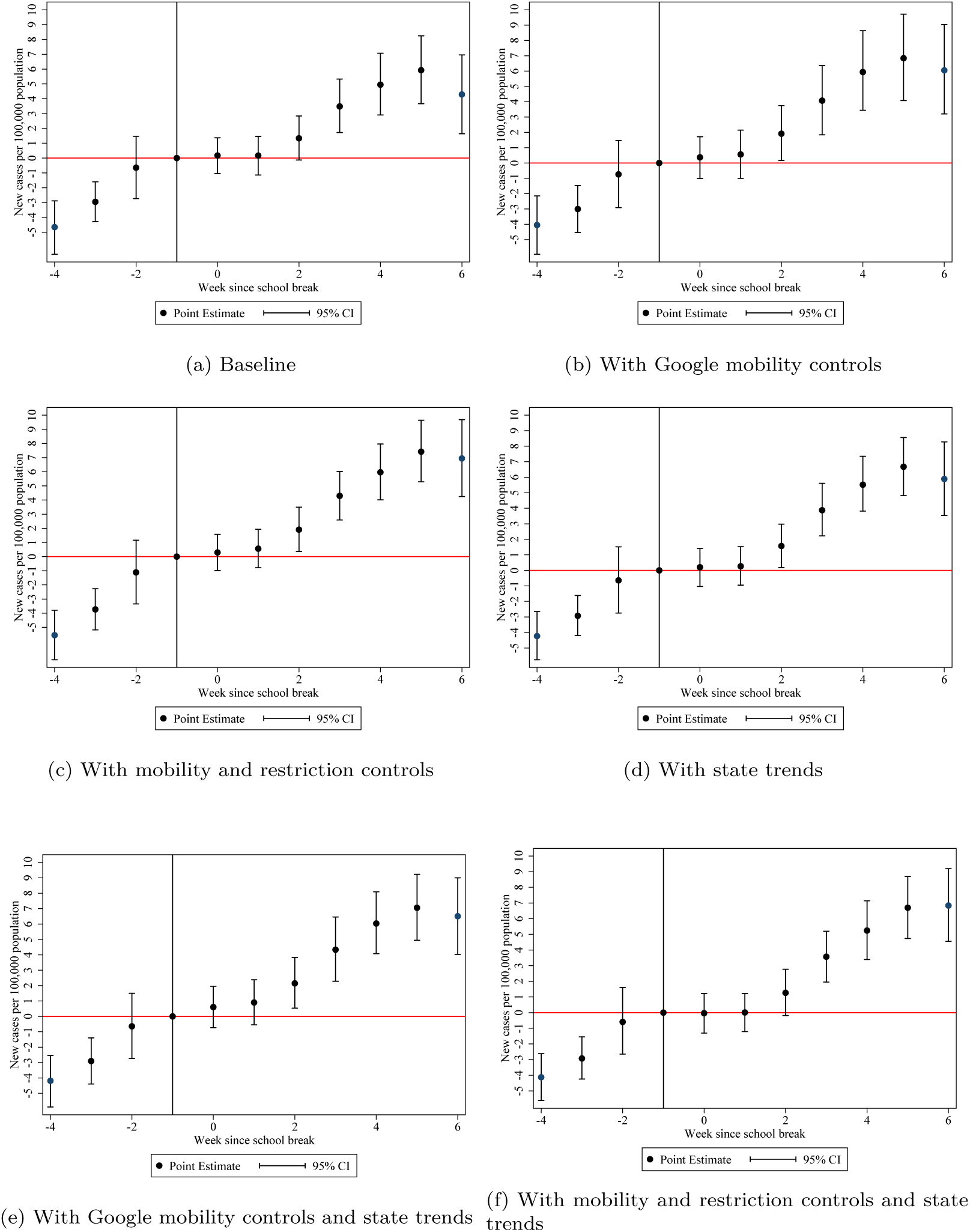
New cases of COVID-19 per 100k population and school breaks in Germany *Notes:* All graphs display the weekly new confirmed cases of COVID-19 infections in Germany. The effects are estimated by binning all weeks beyond the maximum Leads and Lags before and since the beginning of the school breaks. Standard errors are computed by the wild clustered bootstrap method and clustered at the state level.

Panel 10a shows the graphical results from estimating the baseline specification without any controls. The coefficients of the forth and third leads are negative and statistically different from zero, indicating a potential violation of the parallel trends assumption. However, the coefficient of the second lead is then very close to zero and statistically insignificant. The first three weeks of the school breaks do not result in a significant increase in weekly case incidence, as the coefficients of the first three lags are small and insignificant. From the third week since the start of the school breaks onward, the point estimates are positive, increasing from week to week, and statistically significant, implying a positive effect of school breaks on COVID-19 incidence. The magnitude of the effects increases from 1.3 new cases of COVID-19 per 100,000 population for the second lag relative to the baseline period to 5.9 new cases for the fifth lag, followed by a slight decline to 4.3 new cases for the sixth lag. Panel 10b displays the result after adding the state-level Google mobility controls to the regression, while Panel 10c substitutes the Google controls for the county-level mobility and restriction controls. Interestingly, both the state- and the county-level mobility controls increase the magnitude of the effects estimated for the later weeks of the school breaks. A potential explanation for this association is that in accordance with the patterns described in Figure 8 and Figure 9, mobility increased predominantly in states and counties in the north and east of Germany where the COVID-19 incidence remained at a very low level throughout the summer months. This explanation is consistent with the modeling results by Klüsener *et al*. (2020) that there was room for further relaxing NPIs in Germany during the summer months due to the weak dynamics of the pandemic in this period.

Panel 10d shows the results after adding linear state trends to the baseline regression model. The addition of the state trends increases the magnitude of the estimated effects from the third lead onward but does not alter the dynamic pattern. Adding state-level Google mobility controls further increases the magnitudes (Panel 10e). Substituting the state-level Google mobility controls with county-level mobility controls and a county-level indicator of the severity of the COVID-19-related restrictions still results in a slightly larger magnitude of the estimates compared to the baseline regression, while leaving the dynamics of the effects unaffected (Panel 10f). The regression results underlying these graphs are reported in Table 4.

**Table 4:** Baseline estimates

|  | (1) | (2) | (3) | (4) | (5) | (6) |
| --- | --- | --- | --- | --- | --- | --- |
|  | New cases of COVID-19 per 100,000 population |  |  |  |  |  |
| Lead 4 | -4.656***<br>(0.612) | -4.049***<br>(0.631) | -5.554***<br>(0.637) | -4.234***<br>(0.661) | -4.185***<br>(0.701) | -4.129***<br>(0.706) |
| Lead 3 | -2.952***<br>(0.578) | -3.004***<br>(0.592) | -3.731***<br>(0.594) | -2.924***<br>(0.569) | -2.904***<br>(0.597) | -2.930***<br>(0.601) |
| Lead 2 | -0.645<br>(0.551) | -0.741<br>(0.554) | -1.114**<br>(0.551) | -0.643<br>(0.538) | -0.646<br>(0.546) | -0.595<br>(0.547) |
| Lag 0 | 0.174<br>(0.552) | 0.369<br>(0.601) | 0.295<br>(0.554) | 0.202<br>(0.538) | 0.604<br>(0.610) | -0.036<br>(0.545) |
| Lag 1 | 0.169<br>(0.586) | 0.560<br>(0.708) | 0.563<br>(0.600) | 0.266<br>(0.575) | 0.903<br>(0.750) | 0.009<br>(0.603) |
| Lag 2 | 1.333**<br>(0.638) | 1.914**<br>(0.814) | 1.909***<br>(0.672) | 1.572**<br>(0.629) | 2.144**<br>(0.862) | 1.265*<br>(0.684) |
| Lag 3 | 3.482***<br>(0.689) | 4.071***<br>(0.879) | 4.296***<br>(0.740) | 3.875***<br>(0.681) | 4.334***<br>(0.943) | 3.569***<br>(0.760) |
| Lag 4 | 4.949***<br>(0.757) | 5.937***<br>(0.923) | 5.964***<br>(0.825) | 5.520***<br>(0.748) | 6.035***<br>(0.984) | 5.242***<br>(0.844) |
| Lag 5 | 5.924***<br>(0.822) | 6.837***<br>(0.927) | 7.421***<br>(0.909) | 6.678***<br>(0.811) | 7.059***<br>(0.970) | 6.700***<br>(0.930) |
| Lag 6 | 4.291***<br>(0.924) | 6.048***<br>(0.970) | 6.942***<br>(1.048) | 5.889***<br>(0.909) | 6.507***<br>(0.973) | 6.837***<br>(1.056) |
| Observations | 7218 | 7170 | 7218 | 7218 | 7170 | 7218 |
| County FE | Yes | Yes | Yes | Yes | Yes | Yes |
| Week FE | Yes | Yes | Yes | Yes | Yes | Yes |
| Linear state trends | No | No | No | Yes | Yes | Yes |
| Google controls | No | Yes | No | No | Yes | No |
| Mobility controls | No | No | Yes | No | No | Yes |
| Restriction controls | No | No | Yes | No | No | Yes |

The table reports event-study estimates of the effect of the school breaks on the COVID-19 incidence per 100,000 population in Germany. All regression models were estimated by binning weeks beyond the maximum included number of Leads and Lags. The Lead estimates indicate weeks prior to the beginning of the school breaks. The last week before the beginning of the school breaks is omitted. The Lag estimates indicate weeks since the beginning of the school breaks. Column 1 reports results from performing the regression without additional controls besides the county and week FE. Column 2 reports results from adding Google mobility controls to the regression. Column 3 reports results from adding state-level mobility and restriction controls to the regression. Columns 4-6 report results from repeating the regressions reported in columns 1-3 with linear state trends added to the regressions. Bootstrapped standard errors clustered at the state level reported in parentheses. \*\*\* $p < 0.01$ , \*\* $p < 0.05$ , \* $p < 0.1$

The dynamic pattern of initially small effects that increase substantially in magnitude the longer the states are on school breaks is consistent with international travel movements of residents picking up pace with the start of the school breaks and returning travelers importing infections from abroad upon completion of their trips a few weeks later. The initial zero effects of the school breaks are furthermore consistent with the findings by Isphording *et al*. (2020) and von Bismarck-Osten *et al*. (2020) that the school closures have not significantly affected the COVID-19 incidence in Germany. Finally, the fact that the point estimate of the sixth lag is smaller than the point estimate of the earlier fifth lag in most regressions reported here is consistent with the finding of von Bismarck-Osten *et al*. (2020) that also the school reopenings after the school breaks have not caused a surge in COVID-19 incidence.

### 6.2 Robustness

Figure 11 presents a series of robustness checks. Panel 11a displays the coefficients from estimating the baseline regression model with the county-level mobility and restriction controls but omitting the three city states of Berlin, Bremen, and Hamburg from the regression. In Panel 11b, the state-level Google mobility controls are lagged by one week in order to allow for a potentially delayed effect on the COVID-19 incidence, while in Panel 11c, the county-level mobility and restriction controls are lagged. Panels 11d-11f repeat the previous three estimations but with linear state trends added to the regressions. The various modifications primarily affect the precision of the individual estimates but do not change the dynamic pattern. The regression results of the robustness checks are reported in Table 5.

**Figure 11:**
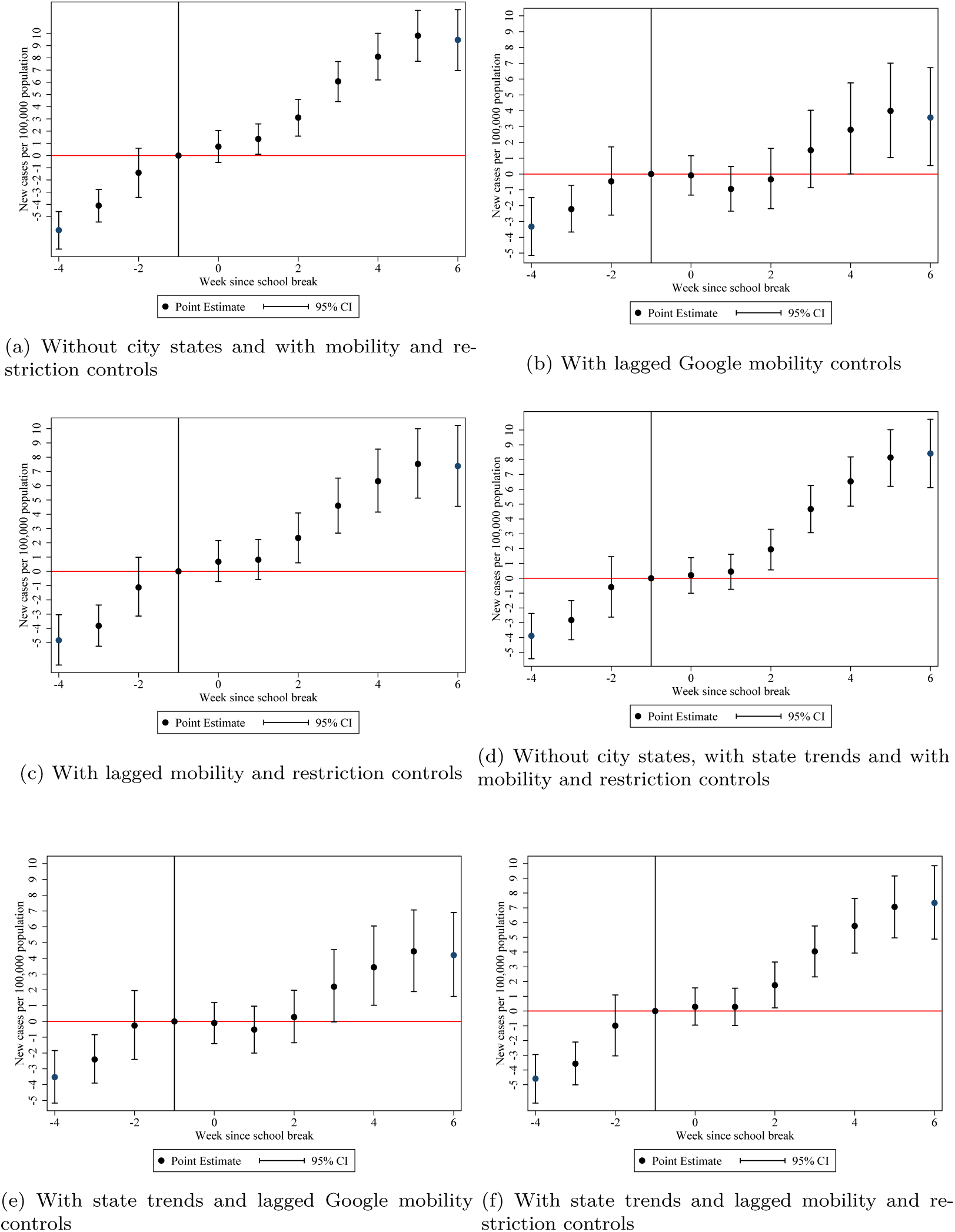
Robustness: New cases of COVID-19 per 100k population and school breaks in Germany *Notes:* All graphs display the weekly new confirmed cases of COVID-19 infections in Germany. The effects are estimated by binning all weeks beyond the maximum Leads and Lags before and since the beginning of the school breaks. Standard errors are computed by the wild clustered bootstrap method and clustered at the state level.

**Table 5:** Robustness checks

|  | (1) | (2) | (3) | (4) | (5) | (6) |
| --- | --- | --- | --- | --- | --- | --- |
|  | New cases of COVID-19 per 100,000 population |  |  |  |  |  |
| Lead 4 | -6.108***<br>(0.666) | -3.324***<br>(0.635) | -4.827***<br>(0.625) | -3.882***<br>(0.743) | -3.529***<br>(0.682) | -4.587***<br>(0.674) |
| Lead 3 | -4.094***<br>(0.623) | -2.217***<br>(0.596) | -3.813***<br>(0.586) | -2.819***<br>(0.636) | -2.406***<br>(0.594) | -3.571***<br>(0.590) |
| Lead 2 | -1.407**<br>(0.574) | -0.464<br>(0.567) | -1.123**<br>(0.553) | -0.597<br>(0.574) | -0.260<br>(0.560) | -0.995*<br>(0.546) |
| Lag 0 | 0.731<br>(0.574) | -0.086<br>(0.556) | 0.679<br>(0.550) | 0.207<br>(0.569) | -0.099<br>(0.546) | 0.291<br>(0.546) |
| Lag 1 | 1.362**<br>(0.621) | -0.946<br>(0.661) | 0.815<br>(0.596) | 0.450<br>(0.626) | -0.514<br>(0.669) | 0.285<br>(0.602) |
| Lag 2 | 3.113***<br>(0.694) | -0.339<br>(0.788) | 2.341***<br>(0.662) | 1.952***<br>(0.707) | 0.277<br>(0.814) | 1.754***<br>(0.680) |
| Lag 3 | 6.072***<br>(0.765) | 1.505*<br>(0.905) | 4.602***<br>(0.732) | 4.669***<br>(0.784) | 2.204**<br>(0.934) | 4.044***<br>(0.758) |
| Lag 4 | 8.096***<br>(0.863) | 2.798***<br>(0.995) | 6.315***<br>(0.815) | 6.531***<br>(0.884) | 3.431***<br>(1.033) | 5.770***<br>(0.848) |
| Lag 5 | 9.816***<br>(0.949) | 3.991***<br>(1.049) | 7.526***<br>(0.892) | 8.144***<br>(0.972) | 4.440***<br>(1.082) | 7.065***<br>(0.923) |
| Lag 6 | 9.466***<br>(1.096) | 3.575***<br>(1.044) | 7.378***<br>(1.015) | 8.417***<br>(1.108) | 4.201***<br>(1.059) | 7.334***<br>(1.041) |
| Observations | 7146 | 7170 | 7218 | 7146 | 7170 | 7218 |
| County FE | Yes | Yes | Yes | Yes | Yes | Yes |
| Week FE | Yes | Yes | Yes | Yes | Yes | Yes |
| Linear state trends | No | No | No | Yes | Yes | Yes |
| City states | No | Yes | Yes | No | Yes | Yes |
| Mobility controls | Yes | No | No | Yes | No | No |
| Restrictions controls | Yes | No | No | Yes | No | No |
| Lagged Google controls | No | Yes | No | No | Yes | No |
| Lagged mobility controls | No | No | Yes | No | No | Yes |
| Lagged restriction controls | No | No | Yes | No | No | Yes |

The table reports event-study estimates of the effect of the school breaks on the COVID-19 incidence per 100,000 population in Germany. All regression models were estimated by binning weeks beyond the maximum included number of Leads and Lags. The Lead estimates indicate weeks prior to the beginning of the school breaks. The last week before the beginning of the school breaks is omitted. The Lag estimates indicate weeks since the beginning of the school breaks. Column 1 reports results from omitting the three German city states from the regression. Column 2 reports results from adding the first lag of the Google mobility controls to the regression. Column 3 reports results from adding the first lag of the county-level mobility and restriction controls to the regression. Columns 4-6 report results from repeating the regressions reported in columns 1-3 with linear state trends added to the regressions. Bootstrapped standard errors clustered at the state level reported in parentheses. \*\*\* $p < 0.01$ , \*\* $p < 0.05$ , \* $p < 0.1$

Instead of binning the lead and lag periods beyond the specified maximum lead and lag periods, the eventdd routine further provides the option of using a balanced panel including only leads and lags up to the specified maximum for each unit of observation. Table 6 reports a set of robustness estimates from using only a balanced panel of counties and weeks. Column 1 reports results from estimating the baseline specification. Column 2 reports results from adding the time-varying county-level mobility and restriction controls. Column 3 reports results from estimating the baseline specification without the three city states. Column 4 reports results from adding the first lag of the mobility and restriction controls to the baseline specification. The corresponding graphical results are displayed in Figure 12. All results are very similar in terms of magnitude, significance, and dynamics to the estimates obtained from applying the binning approach.

**Figure 12:**
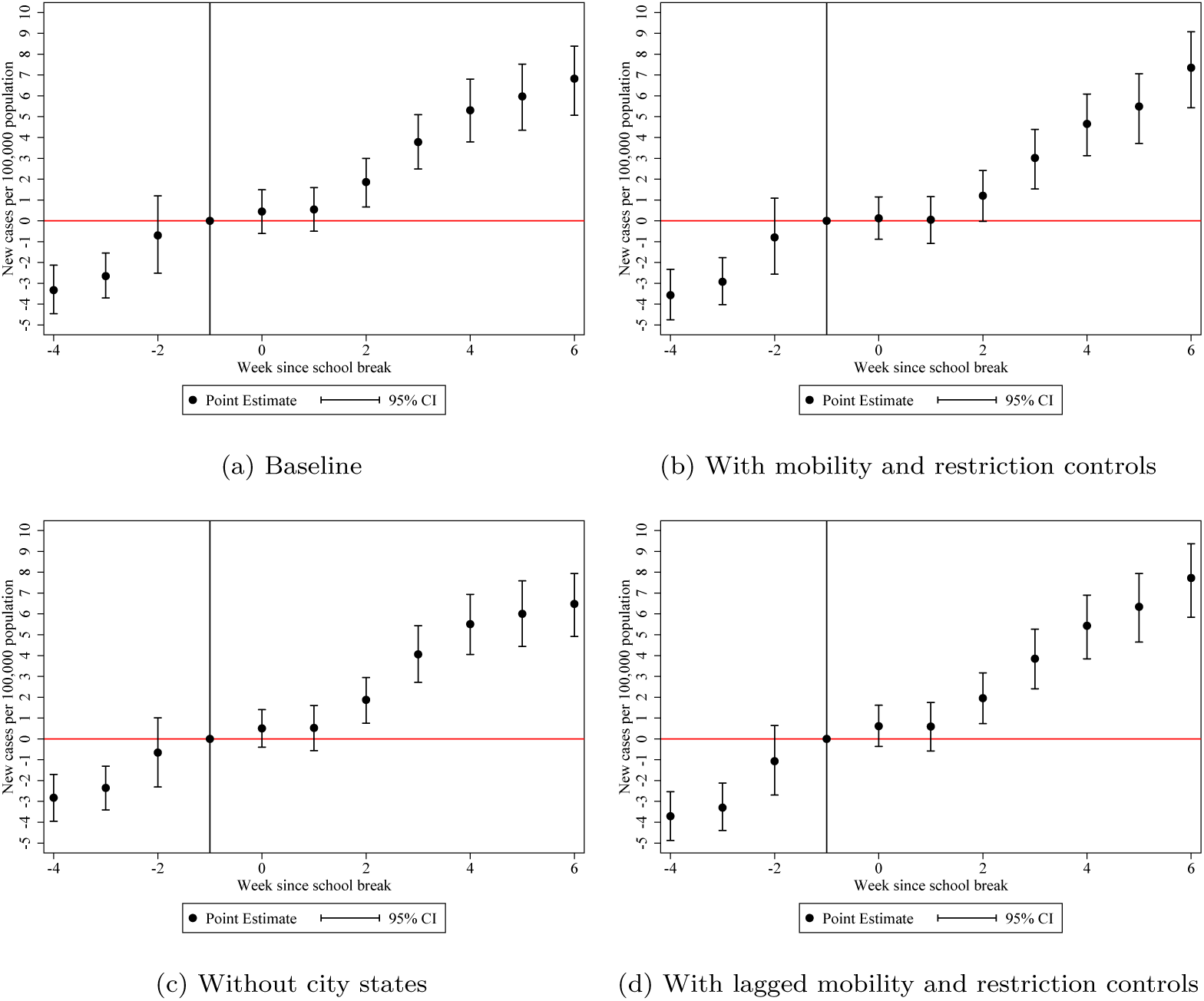
Robustness: New cases of COVID-19 per 100k population and school breaks in Germany *Notes:* All graphs display the weekly new confirmed cases of COVID-19 infections in Germany. The effects are estimated using a balanced panel of weeks before and since the beginning of the school breaks. Standard errors are computed by the wild clustered bootstrap method and clustered at the state level.

**Table 6:**
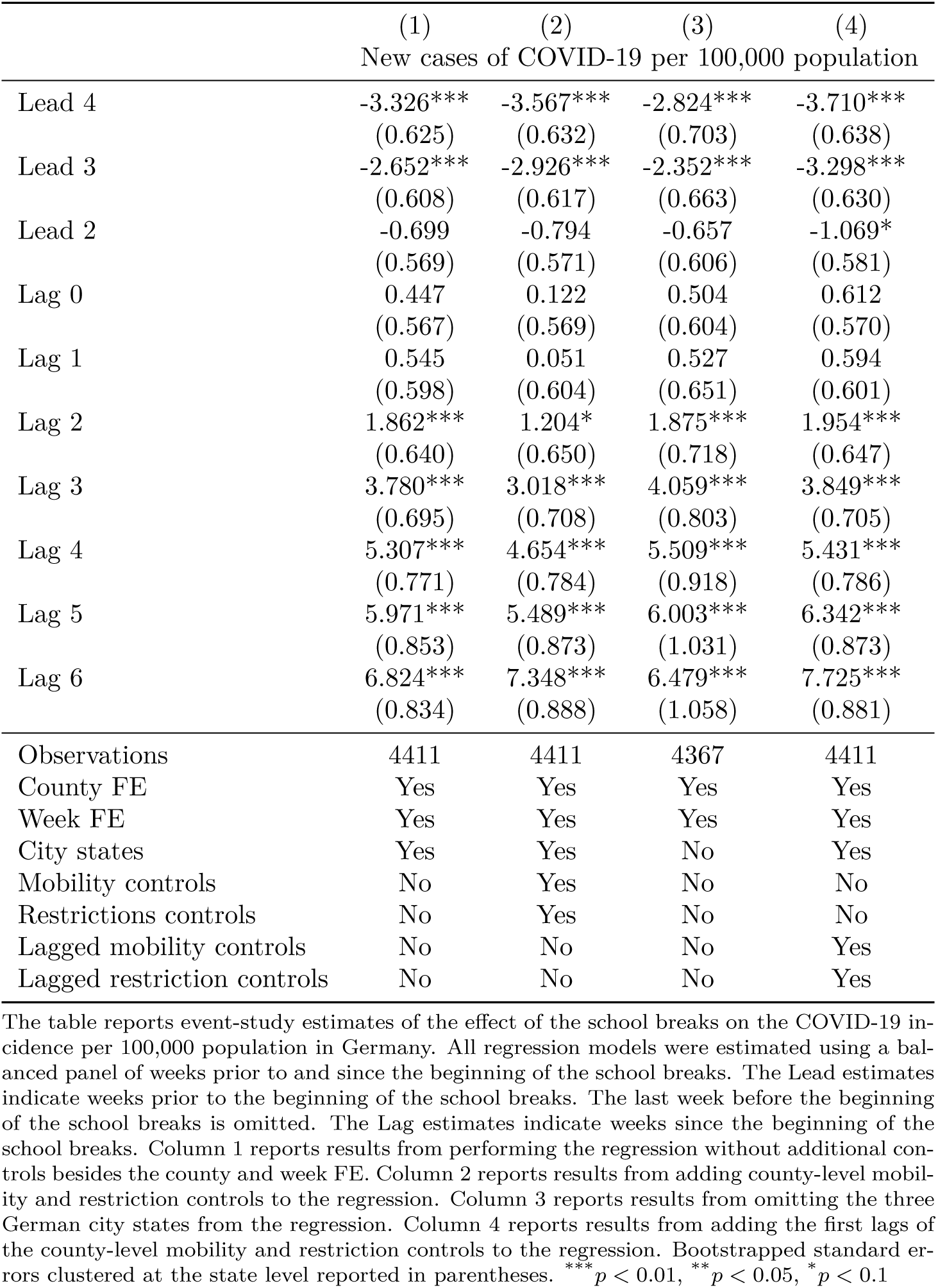
Balanced panel estimates

Furthermore, the main regression including the state trends and the mobility and restriction controls is repeated 16 times while omitting a different state each time from the regression. The graphs and the table containing the results of this exercise are relegated to the Appendix. Figure A1 and Table A1 indicate that the main results are not driven by one particular state. However, omitting North Rhine-Westphalia considerably decreases the magnitude of the negative estimates of the fourth and third leads. This effect might be due to a number of large outbreaks of COVID-19 in meat-processing plants in various counties of North Rhine-Westphalia during May and June.

### 6.3 Testing

Recall that free tests for travel returnees were offered since August 1, while the mandatory free testing regime for travel returnees from risk areas was introduced on August 8. Realigning the testing capacities towards the travel returnees has certainly improved the surveillance of imported infections. However, this particular focus raises the question whether the estimates indicating a significant increase in incidence during the later weeks of the school breaks are partly driven not only by travelers being at greater risk of infection but also facing a greater chance of being detected as infected upon return than the non-traveling population.

First, there is little reason to suspect that the realignment of testing capacities towards the travel returnees has compromised the detection of cases among the non-traveling population in Germany. A representative seroprevalence survey conducted in the city of Munich estimates that the ratio of undetected infections to confirmed cases (German: *Dunkelziffer*) was less than two during the summer months; a decline from a ratio of four during the first wave in spring (LMU Munich University Hospital Division of Infectious Diseases and Tropical Medicine and HelmholtzZentrum Munich, 2020). While this finding does not necessarily generalize beyond Munich, it corresponds to the very low share of COVID-19 tests that were positive in Germany during the summer months.

Second, the overlap of the school breaks with at least one of the two testing regimes was quite heterogeneous across states: One state’s school breaks ended on August 1 (Mecklenburg-Vorpommern). Two more states concluded their breaks before August 8 (Berlin and Hamburg), while two other states concluded them exactly on August 8 (Brandenburg and Schleswig-Holstein). Four states’ school breaks ended within the first week following the start of the mandatory testing regime (Hesse, North Rhine-Westphalia, Rhineland-Palatinate, Saarland). A remainder of five states spent at least the final two weeks of their school breaks under both testing regimes (Bremen, Lower Saxony, Saxony, Saxony-Anhalt, Thuringia). Finally, two states began their school breaks less than one week before the start of the free testing regime and less than two weeks before the start of the mandatory testing regime (Baden-Württemberg and Bavaria). The robustness checks in Table A1 and Figure A1 indicate that the estimated effects rather increase than decrease if one of the latter two states is excluded from the regressions, suggesting they are not driving the results.

Third, the introduction of the two testing regimes coincided with a number of factors that are similarly inclined to drive up the COVID-19 incidence among returning travelers: (1) The absolute number of travel returnees was increasing due to the nearing end of the school breaks in several states, which should *ceteris paribus* increase the number of cases found among travel returnees. (2) The COVID-19 incidence in several important travel destinations was growing, which should *ceteris paribus* increase the number of cases found among travel returnees. (3) The number of countries designated as risk areas was growing, which increased the scope of the mandatory testing regime, thereby increasing also the potential for ‘over’-testing the travel returnees by shifting them from the voluntary to the mandatory regime. (4) The composition of risk areas from which travelers have been returning to Germany has been changing throughout the summer months, whereas it is difficult to assess how the infection risk has changed within the group of risk areas.

The general complexity of the testing situation is underlined by Figure 13. The left panel shows the weekly incidence of COVID-19 in Germany, differentiating between cases with likely place of infection within Germany and cases with likely place of infection abroad. The right panel performs the same differentiation regarding the share of cases that is attributed to each of the two potential locations of infection. Both the incidence and the share of cases from abroad increase markedly with the introduction of the free testing regime in week 32 and the mandatory testing regime in week 33. However, the two variables show considerable variation while both testing regimes were in place, with both variables declining already well before all states have completed their school breaks and the free testing regime has been terminated in week 37. Figure A2 in the Appendix differentiates the two places of likely infection further by state, with the dashed line indicating if the mandatory testing regime was in place. The patterns across states are heterogeneous: In some states, the mandatory testing regime was associated with a sharp rise in cases with place of infection abroad, while the latter have already been on the rise in other states before the introduction of the mandatory regime. The cases in the states in east Germany whose school breaks at least partially overlapped with the mandatory testing regime hardly showed a reaction.

**Figure 13:**
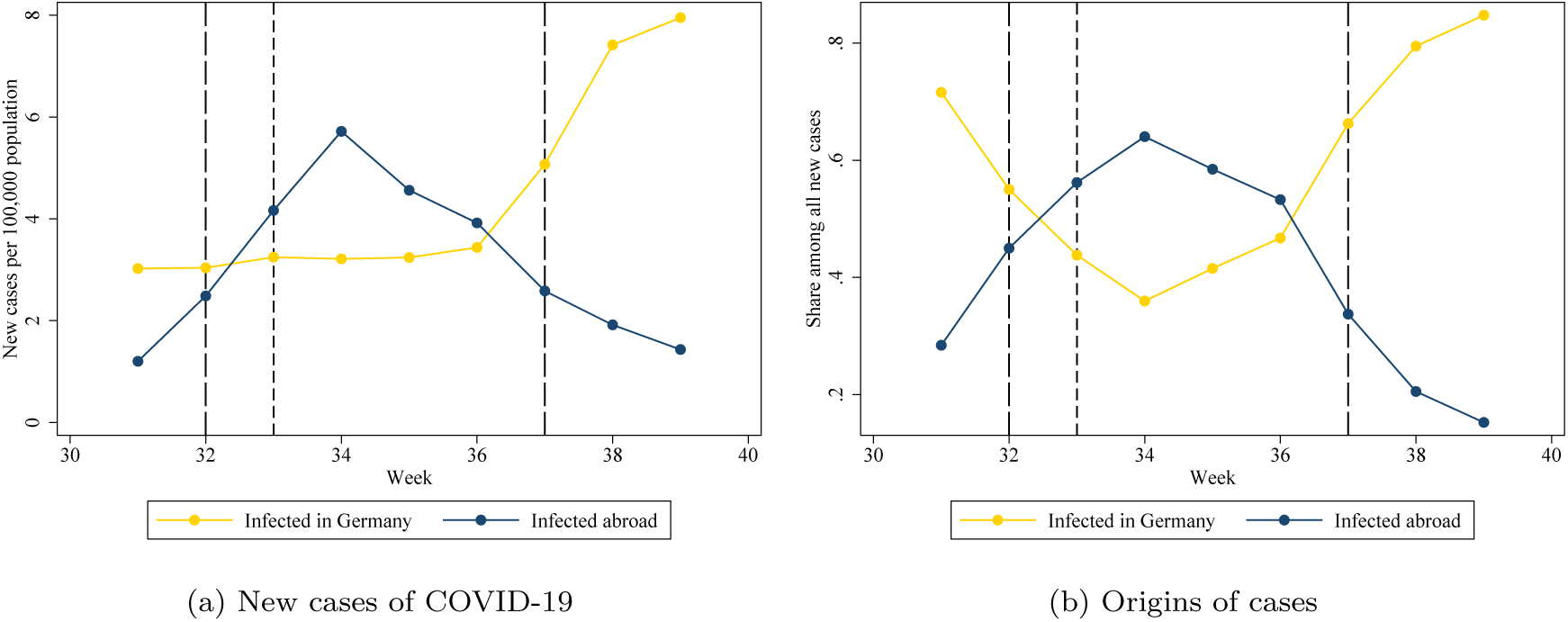
Cases of COVID-19 in Germany differentiated by places of infection *Notes:* Source: Various RKI (2020) situation reports. The left panel shows the weekly incidence of COVID-19 per 100,000 population in Germany. The golden line indicates the incidence of cases with likely infection in Germany. The blue line indicates the incidence of cases with likely infection abroad. The two vertical red lines indicate the first and the last weeks of the school breaks respectively. The vertical short-dashed black line indicates the start of the voluntary testing regime for travelers returning from non-risk areas. The vertical dashed black line indicates the start of the mandatory testing regime for travelers returning from declared risk areas.

Hence, while the particular testing focus on travel returnees during the summer months has likely increased the incidence among travel returnees by a certain factor relative to the non-traveling population, attributing the rising incidence entirely to testing seems premature.

## 7 Discussion

First, it is worth pointing out again the context and setting of this study: The restrictions on international travel were loosened during a period of low COVID-19 incidence in Germany, while the COVID-19 incidence in other European countries was fluctuating considerably. This setting facilitates the detection of a rising COVID-19 incidence due to international travel, as it suggests a rather straight-forward mechanism from exposure and infection abroad to detection and potential secondary infections upon return to Germany.

Interpreting the rising incidence in Germany during school breaks as the effect of revived international mobility without being able to distinguish between the traveling and the non-traveling residents would be difficult to justify in most empirical settings. Within the context of the summer months of 2020, the justification of this interpretation rests on (1) the low incidence in Germany prior and at the beginning of the school breaks, (2) the precise controls for mobility and COVID-19-related restrictions within Germany, and (3) the descriptive associations between the travel movements to countries with higher incidences as well as on the detected cases among infected returnees.

Second, a number of potential sources for bias in the estimated effects can be considered, as suggested by Goodman-Bacon and Marcus (2020): The empirical approach of this paper implicitly assumes that the cases of infected travel returnees are registered in their respective state of residence; with an identical assumption applying to secondary infections caused by travel returnees. While there is little reason to question the first assumption, the second one might be more questionable if infected travel returnees traveled extensively across state borders following their return to Germany. If these domestic movements by infected travel returnees occurred and if they caused significant infections in states other than the respective returnees’ state of residence, these spillovers of infections would downward-bias the estimates if the states affected by the spillovers were not on school break yet. If, in turn, they were already on school break, too, but started in a different calendar week, the spillovers would only affect the relative magnitude of the effects. Finally, if they were on school break and if they went on school break at the same time, the spillovers would not bias the estimates, as the spillovers would occur among the group of treated states.

In addition, it is worth noting that Germany received not only an influx of travel returnees with residence in Germany but also an influx of tourists from abroad during the summer months. Some of these tourists might have been infected with SARS-CoV-2, thereby carrying the risk of causing further infections among residents of Germany to which they have been in contact with during their stay. During the four months from June to September, accommodation providers in Germany reported 5.7 million arrivals of guests from abroad (Statistisches Bundesamt (Destatis), 2020d). However, 4.2 million of these guests arrived from neighboring states of Germany where the COVID-19 incidence had been fairly low until close to the end of September. Infections introduced into Germany from abroad by foreign tourists have not brought up as a concern by the RKI during the same period. Further, the incidence of these potential cases would have to be aligned with the timings of the German school breaks in order to bias the results, for which there is no obvious reason.

Third, recall that the estimates presented in this study should be interpreted as intention-to-treat (ITT) effects, as not every resident in Germany experienced a shock to her/his opportunities to travel from the combination of the relaxed restrictions and the onset of the school breaks - and among those who did, not all have actually traveled abroad during summer. This partial non-compliance dilutes the magnitude of the effects. Further, two important factors affecting the generalizability of the ITT effects are the characteristics of the travelers and the choice of the travel destination countries. According to RKI reports, a large share of infected travelers returned from the Southeast European countries of Kosovo and Bosnia and Herzegovina, which are not popular destinations among German tourists. The RKI situation reports suggests that family visits were the main motivation for travelers from Germany towards these countries. Continuing along this notion, family visits likely involve much closer contact of the travelers to the local population than touristic trips do. Hence, travelers to Southeast European countries not only traveled to countries with a relatively high incidence but also faced a potentially higher risk of infection due to their contact behavior there. If these particular travelers were furthermore more inclined to travel for the purpose of visiting their families than people who would travel only for touristic purposes, then this would render the ITT effects rather specific, as the traveler composition of the summer months would not be representative of the population that had the opportunity to travel. However, the RKI also reported a high number of young infected returning travelers who had traveled for touristic purposes to destinations such as Croatia.

Fourth, this study did not consider whether the loosened travel restrictions may also have affected the incidence of deaths related to COVID-19 in Germany. The death toll from COVID-19 has been very low in Germany during the summer months of 2020, which is explained by the relatively young age of the confirmed cases during the same period. While it cannot be ruled out that the infected returning travelers have experienced a severe, potentially fatal course of the disease, or that infected returning travelers have infected other residents in Germany that have died, the low overall death toll during summer does not warrant a statistical association to the travel patterns. Note further that even the highest levels of COVID-19 incidence observed in other European countries during the summer of 2020 have been dwarfed by the incidences observed during the subsequent autumn and winter waves. Consequently, the risk for a traveler of getting infected while abroad has likely increased, too. The estimates reported in this study are therefore not immediately transferable to other epidemiological contexts.

Finally, the estimates from this study can be used for a simple back-on-the-envelope calculation of the implied total incidence of COVID-19 during the period of the summer breaks. The latter can be calculated by multiplying the statistically significant coefficients from each model in Table 4 by Germany’s total population divided by 100,000 and summing up the results for each model. These calculations yield a range from 16,550 to 22,039 confirmed cases, with the model that uses the county-level restriction and mobility controls yielding the upper limit. It could be expected that the total incidence implied by the estimates is somewhat lower than the approx. 23,000 total confirmed cases among travel returnees from the same period due to the ITT character of the estimates. However, given that the estimates presented here would also reflect potential secondary infections caused by travel returnees, this back-on-the-envelope calculation may suggest that the surveillance of travel returnees has been adequate in terms of limiting the risk of creating sustained epidemic dynamics from the imported infections.

## 8 Conclusions

International travel represents an important element of modern lifestyle and a cornerstone to many tourism-oriented economies across the globe. During a pandemic, however, the benefits of international travel, such as the consumer expenditures flowing into the tourism industry, the leisure time enjoyed during vacations, and the important non-pecuniary benefit of visiting family members who live abroad, should be balanced against the public health risks from international mobility.

This study provides evidence that partially reviving international travel during the summer months of 2020 led to a significant increase in the COVID-19 incidence in Germany. In this context, travel took place between a country with a then low incidence, Germany, and several other European countries with higher and more volatile incidence rates. To mitigate the risks from international travel, Germany implemented first a voluntary testing regime for travel returnees and then shortly after a mandatory testing regime for travelers returning from designated risk areas. While this study cannot fully disentangle the effects of the two testing regimes on the incidence among travel returnees from other important time-varying epidemiological factors, it does not find evidence that the public health surveillance has missed a significant number of imported infections or that imported infections have caused a significant number of secondary infections within Germany.

Importantly, several characteristics of this study’s setting do not carry over to other contexts of international travel during the COVID-19 pandemic. Factors such as seasonality and new, potentially more contagious variants of SARS-CoV-2 are suited to make infections among travelers more likely to occur and to increase the risk of importing these infections upon return. Further, significant testing capacities could be realigned towards the travel returnees during the summer months only because the incidence and hence the demand for tests were low within Germany at the same time. Other periods may not allow such a shift but require a significant expansion of testing capacities overall.

## Data Availability

The data on COVID-19 incidence at the county-level in Germany are publicly available at the COVID-19 data hub.
The Google mobility data are publicly available via the Google website.
The data on COVID-19-related restrictions in Germany are available at the Corona Datenplattform after registration.
The data on mobility within Germany are publicly available at the website of the Federal Statistical Office.
The OurWorldInData data are publicly available at the OurWorldInData website.
The population data on Germany are publicly available at the Genesis data hub of the Federal Statistical Office.
The data on COVID-19 incidence at the German state level differentiated by likely place of infection have been provided on request by the Robert Koch-Institute.

https://npgeo-corona-npgeo-de.hub.arcgis.com/

https://www.google.com/covid19/mobility/

https://www.corona-datenplattform.de//

https://www.destatis.de/DE/Service/EXDAT/Datensaetze/mobilitaetsindikatoren-mobilfunkdaten.html

https://ourworldindata.org/coronavirus-data-explorer

https://www-genesis.destatis.de/genesis/online

## Appendices

**Table A1:**
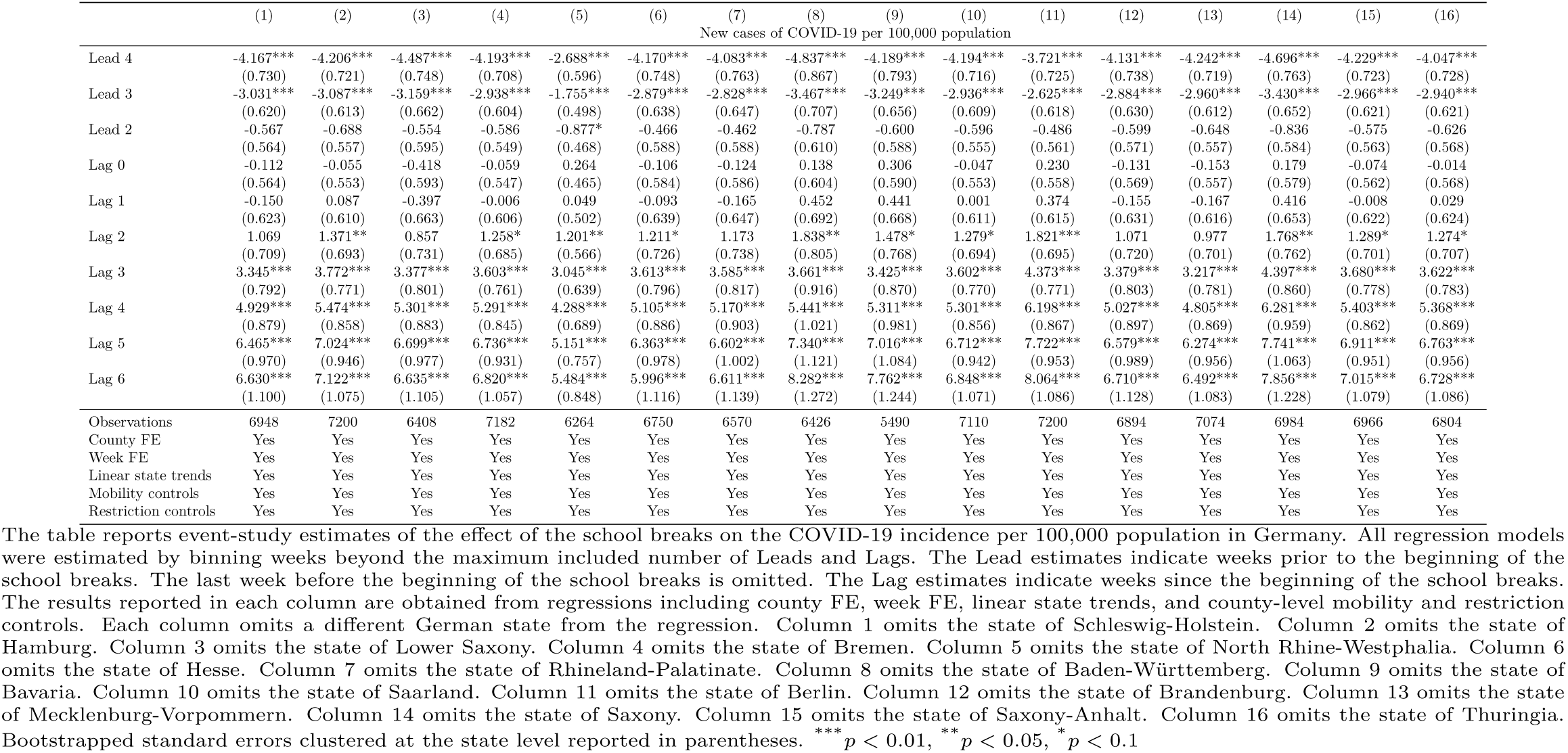
Sensitivity checks

**Figure A1:**
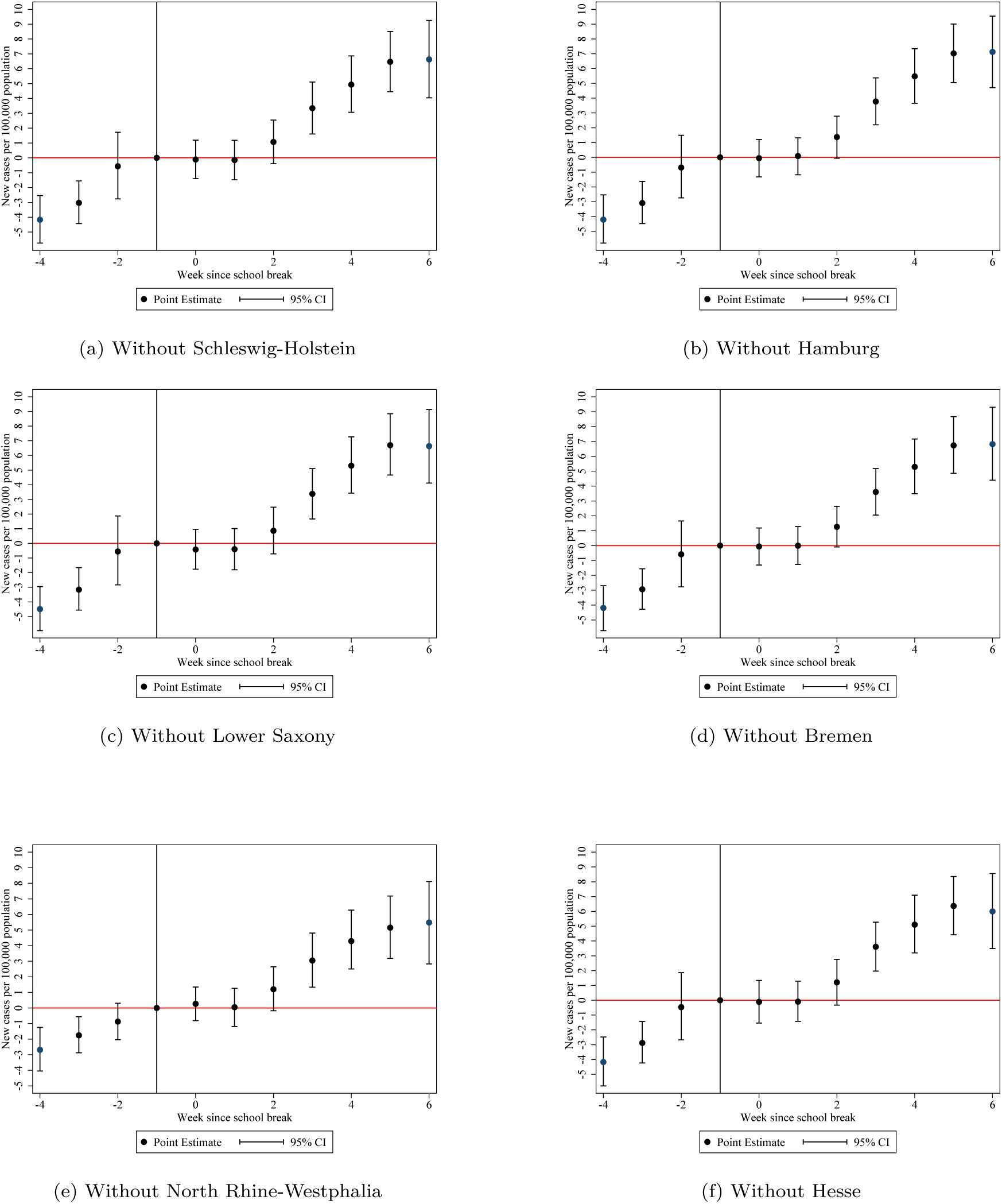
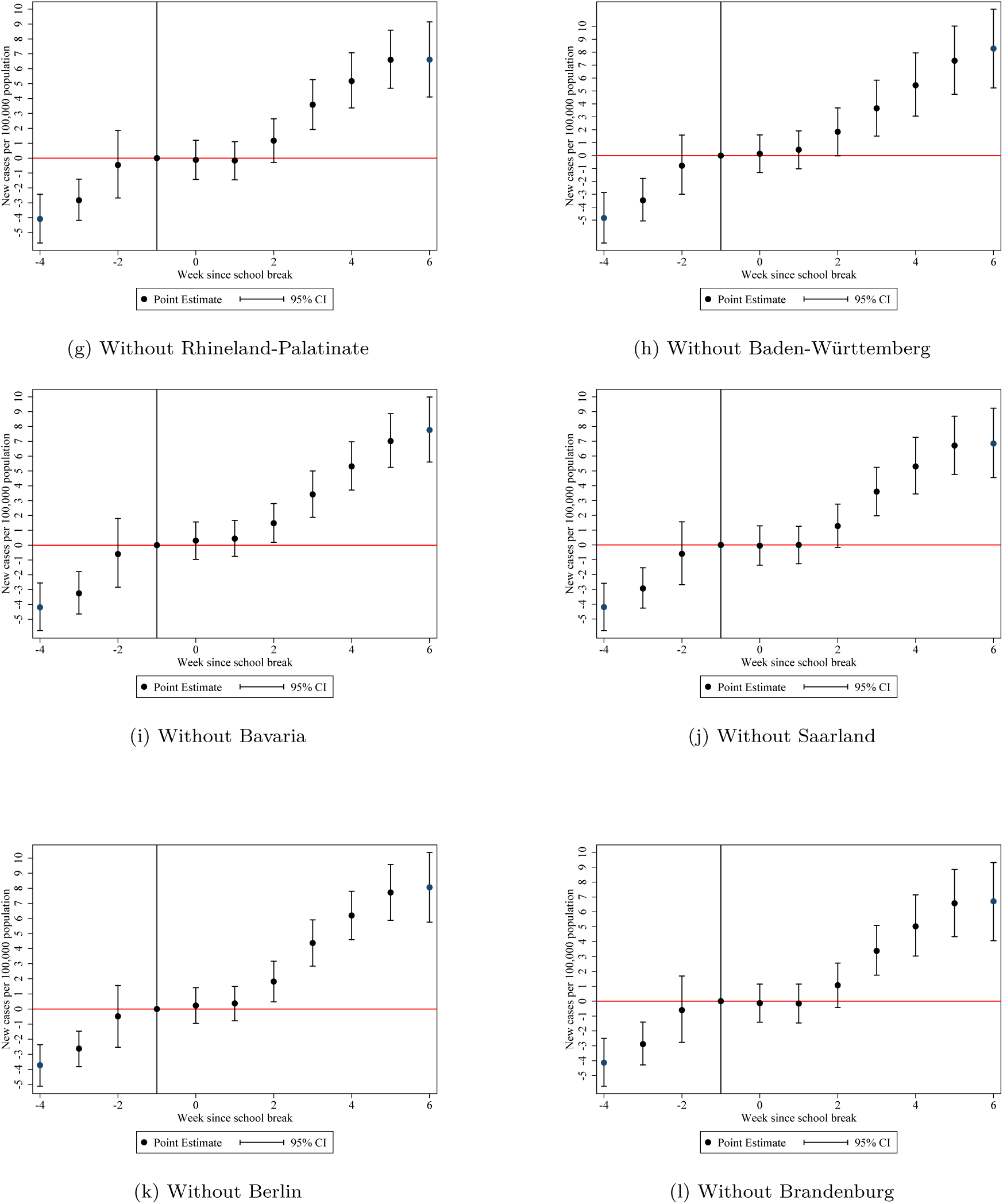
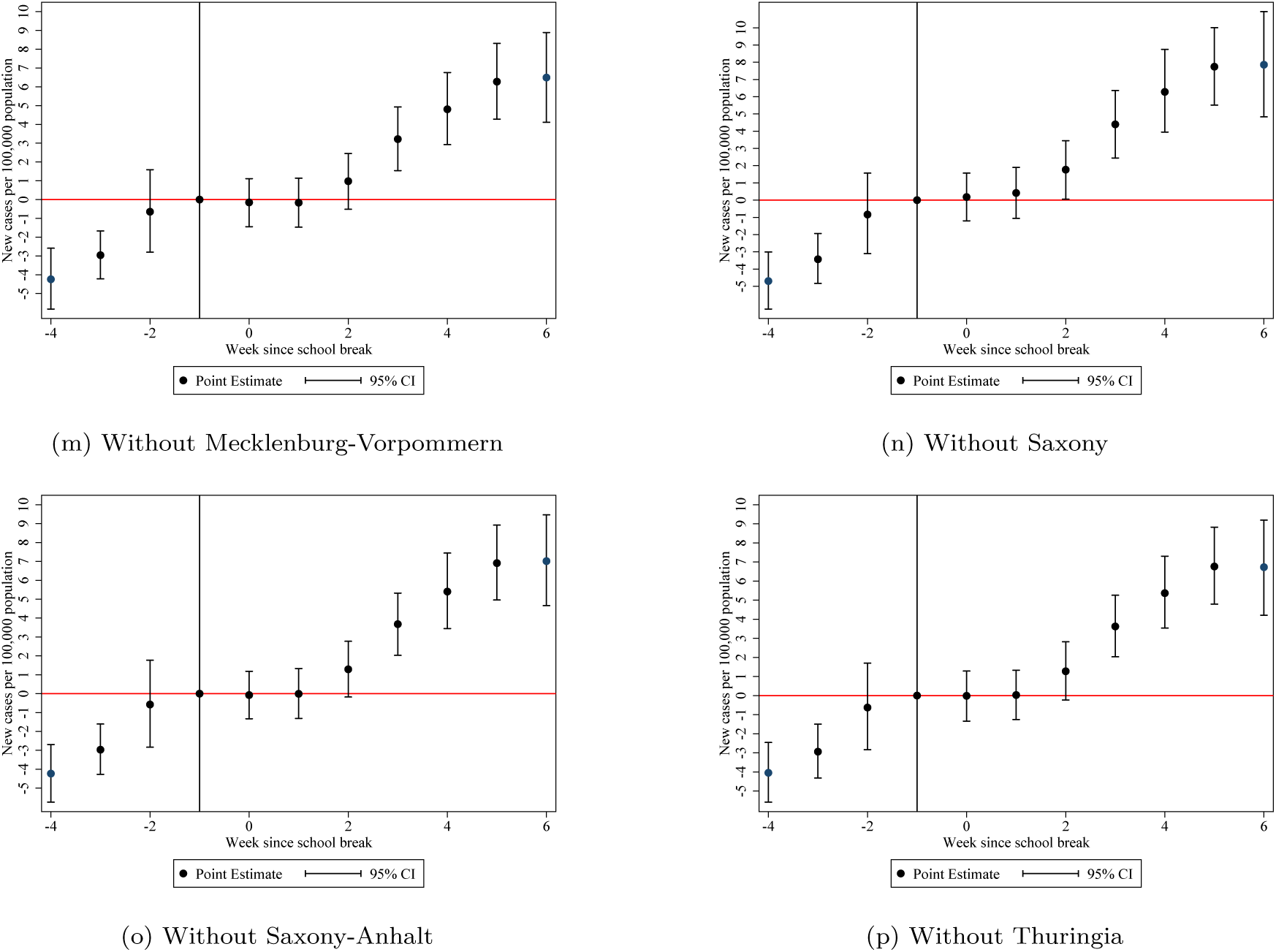
New cases of COVID-19 per 100,000 population and school breaks in German federal states *Notes:* Each graph displays the estimates of new confirmed cases of COVID-19 per 100,000 population in the 16 states of Germany excluding the one indicated in the caption of each graph. The effects are estimated by binning all weeks beyond the maximum Leads and Lags before and since the beginning of the school breaks. Standard errors are computed by the wild clustered bootstrap method and clustered at the state level.

**Figure A2:**
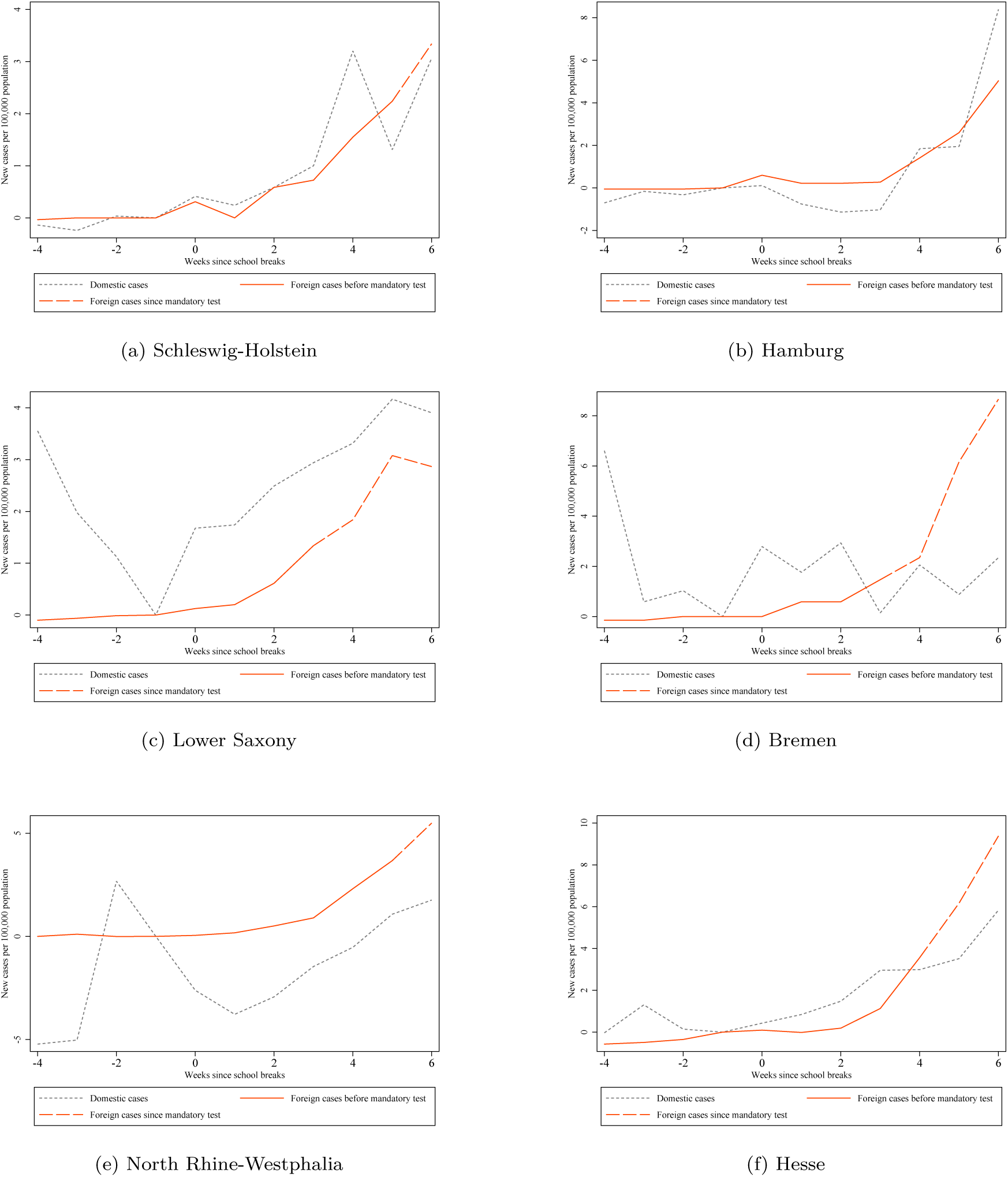
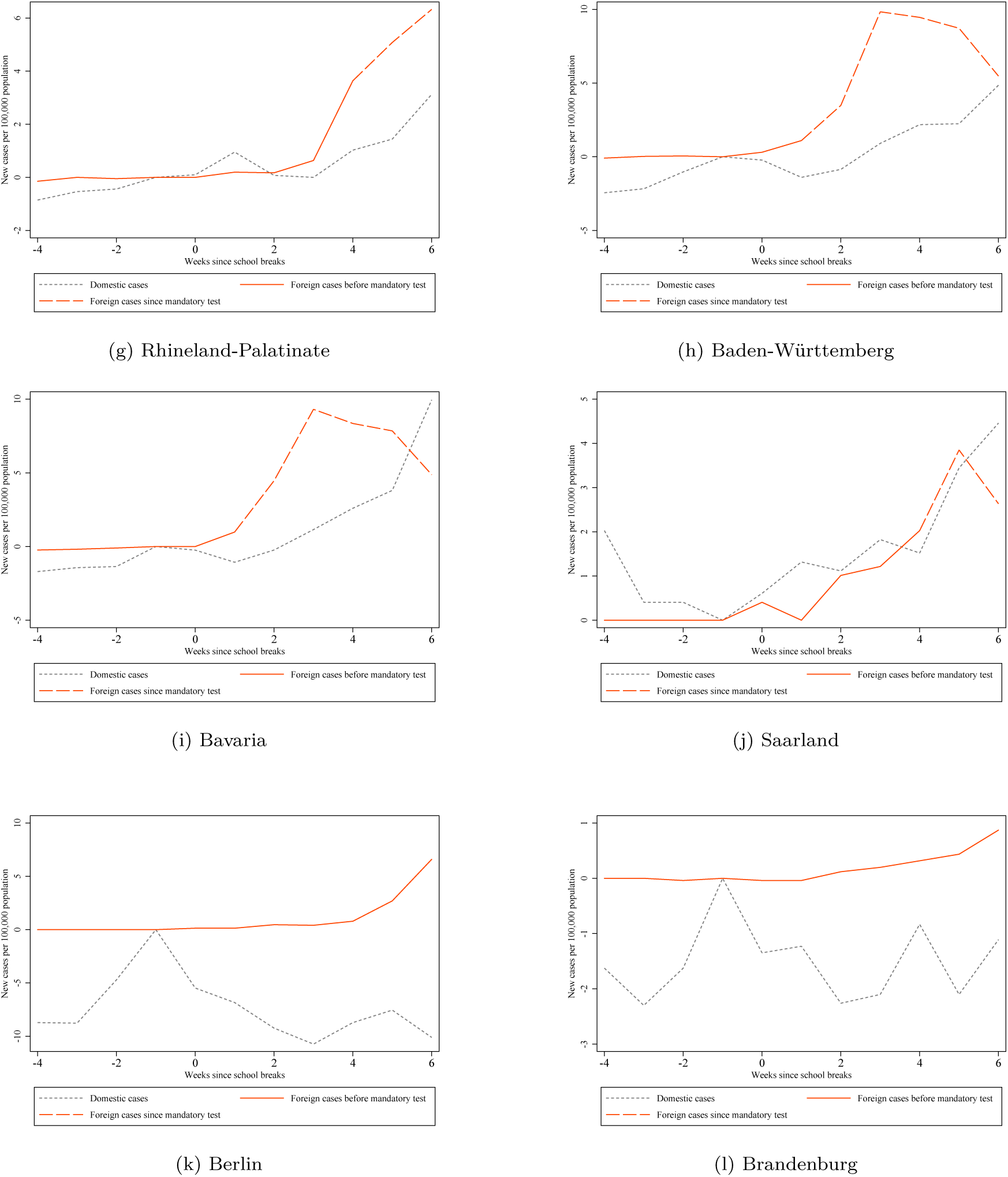
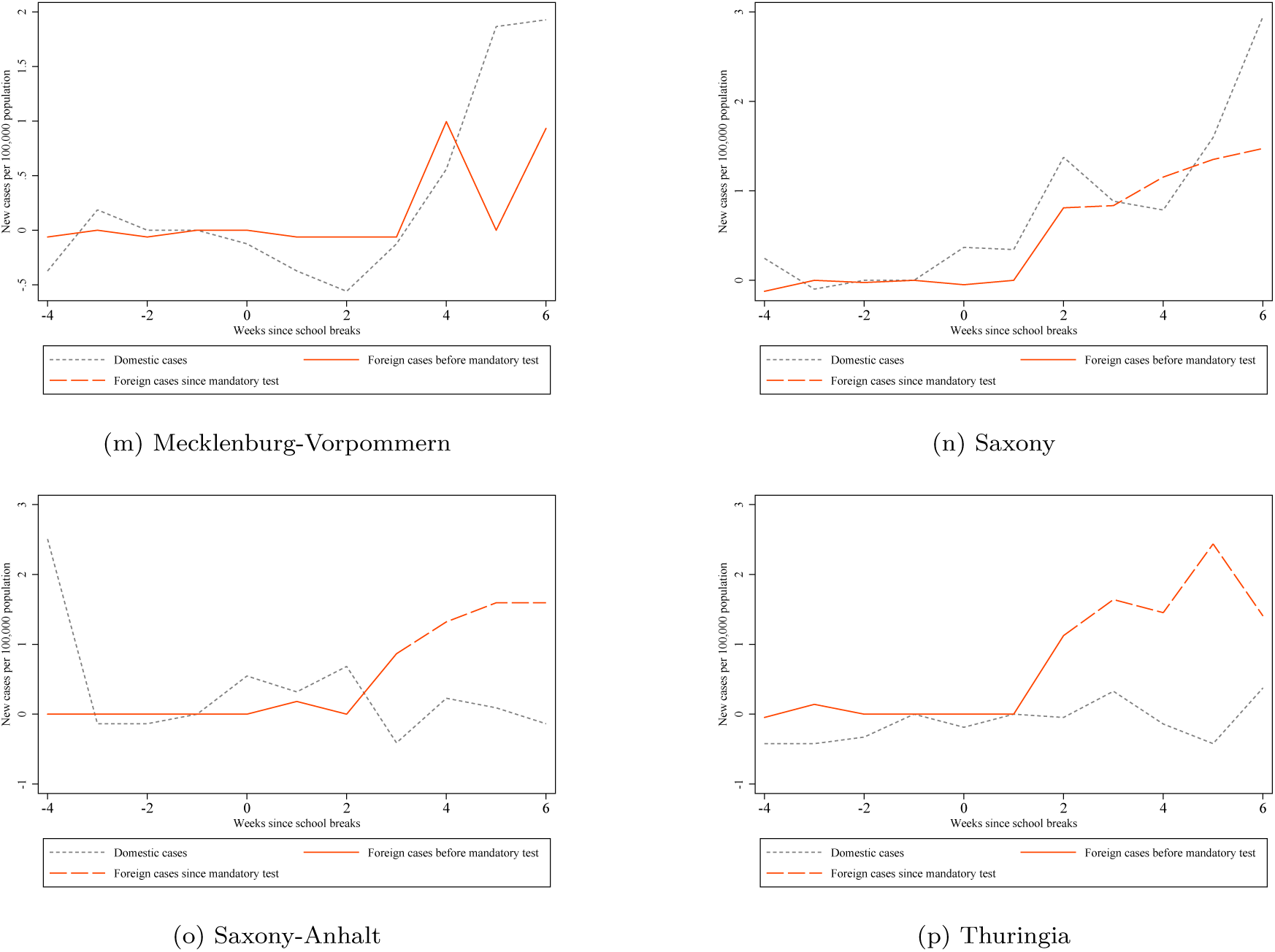
COVID-19 cases in German federal states by place of likely infection *Notes:* Source: RKI (2021). Each graph displays the weekly new confirmed cases of COVID-19 per 100,000 in a German state. The gray line indicates the cases with likely place of infection within Germany. The orange-red line indicates the cases with likely place of infection abroad. The dashed pattern indicates that tests for travelers returning from designated risk areas were mandatory during the respective week.

## Footnotes

1 Robert Koch Institute (RKI), Germany’s federal disease control and prevention agency.

